# Human genetics implicates a BACH2-NRF2 axis in fetal heamoglobin activation

**DOI:** 10.1101/2023.03.24.23287659

**Authors:** Chun-Jie Guo, Uma P. Arora, Xiaoheng Cheng, Wanying Xu, Liam D. Cato, Rick Li, Henry Y. Lu, Andrew J. Lee, Fulong Yu, Gaurav Agarwal, Peng Lyu, Tianyi Ye, Mateusz Antoszewski, Mariel Wissmann, Baraka S. Mkumbe, Supachai Ekwattanakit, Patrick Deelen, Liberata Mwita, Raphael Sangeda, Thidarat Suksangpleng, Suchada Riolueang, Paola G. Bronson, Dirk S. Paul, Emily Kawabata, William J. Astle, Francois Aguet, Kristin Ardlie, Aitzkoa Lopez de Lapuente Portilla, Guolian Kang, Yingze Zhang, Seyed Mehdi Nouraie, Victor R. Gordeuk, Mark T. Gladwin, Melanie E. Garrett, Allison Ashley-Koch, Marilyn J. Telen, Brian Custer, Shannon Kelly, Carla Luana Dinardo, Ester C. Sabino, Paula Loureiro, Anna Bárbara Carneiro-Proietti, Cláudia Maximo, Adriana Méndez, Angelika Hammerer-Lercher, Julie Makani, Vivien A. Sheehan, Mitchell J. Weiss, Lude Franke, Björn Nilsson, Adam S. Butterworth, Vip Viprakasit, Siana Nkya, Vijay G. Sankaran

**Affiliations:** Division of Hematology/Oncology, Boston Children’s Hospital, Harvard Medical School, Boston, Massachusetts, USA; Department of Pediatric Oncology, Dana-Farber Cancer Institute, Harvard Medical School, Boston, Massachusetts, USA; Howard Hughes Medical Institute, Boston, MA 02115, USA; Broad Institute of MIT and Harvard, Cambridge, Massachusetts, USA; Sickle Cell Program, Department of Hematology and Blood Transfusion, Muhimbili University of Health and Allied Sciences, Dar es Salaam, Tanzania; Department of Biochemistry, Muhimbili University of Health and Allied Science, Dar es Salaam, Tanzania; Department of Artificial Intelligence and Innovative Medicine, Graduate School of Medicine, Tohoku University, Sendai, Japan; Siriraj Thalassemia Center, Faculty of Medicine Siriraj Hospital, Mahidol University, Bangkok, Thailand; Department of Genetics, University of Groningen, University Medical Center Groningen, Groningen, the Netherlands; Oncode Institute, Amsterdam, the Netherlands; Department of Pharmaceutical Microbiology, Muhimbili University of Health and Allied Sciences, Dar es Salaam, Tanzania; R&D Translational Biology, Biogen, Cambridge, Massachusetts, USA; British Heart Foundation Cardiovascular Epidemiology Unit, Department of Public Health and Primary Care, University of Cambridge, Cambridge, UK; British Heart Foundation Centre of Research Excellence, University of Cambridge, Cambridge, UK; National Institute for Health and Care Research Blood and Transplant Research Unit in Donor Health and Behaviour, University of Cambridge, Cambridge, UK; MRC Biostatistics Unit, University of Cambridge, Cambridge, UK; NHS Blood and Transplant, Cambridge, UK; Lund Stem Cell Center, Lund University, 221 84 Lund, Sweden; Department of Laboratory Medicine, Lund University, 221 84 Lund, Sweden; St. Jude Children’s Research Hospital, Memphis, Tennessee, USA; Department of Medicine, School of Medicine, University of Pittsburgh, Pittsburgh, Pennsylvania, USA; Division of Hematology and Oncology, Department of Medicine, Comprehensive Sickle Cell Center, University of Illinois at Chicago, Chicago, Illinois, USA; Department of Medicine, University of Maryland School of Medicine, Baltimore, Maryland, USA; Department of Medicine, Duke University Medical Center, Durham, North Carolina, USA; Vitalant Research Institute, San Francisco, California, USA; Department of Laboratory Medicine, UCSF, San Francisco, California, USA; Division of Pediatric Hematology, UCSF Benioff Children’s Hospital, Oakland, California, USA; Fundacao Pro-Sangue Hemocentro de Sao Paulo, Sao Paulo, Brazil; Institute of Tropical Medicine, Faculdade de Medicina da Universidade de Sao Paulo, Sao Paulo, Brazil; Fundacao Hemope, Recife, Pernambuco, Brazil; Fundacao Hemominas, Belo Horizonte, Brazil; Fundacao Hemorio, Rio de Janeiro, Brazil; Institute of Laboratory Medicine, Cantonal Hospital Aarau, 5000 Aarau, Switzerland; Aflac Cancer & Blood Disorders Center, Children’s Healthcare of Atlanta & Department of Pediatrics, Emory University School of Medicine, Atlanta, Georgia, USA; Health Data Research UK Cambridge, Wellcome Genome Campus and University of Cambridge, Cambridge, UK; Heart and Lung Research Institute, University of Cambridge, Cambridge, UK; Department of Pediatrics, Faculty of Medicine Siriraj Hospital, Mahidol University, Bangkok, Thailand; Tanzania Human Genetics Organisation, Dar es Salaam, Tanzania; Harvard Stem Cell Institute, Cambridge, Massachusetts, USA

## Abstract

Human genetic studies have identified key regulators of fetal hemoglobin (HbF) expression, including BCL11A, resulting in therapeutic advances^1–8^. Yet the mechanisms by which HbF expression is activated remain incompletely understood^9^. Here, we conduct a large multi-ancestry genome-wide association study of HbF levels in 28,279 individuals that identifies 91 conditionally-independent associations across 12 genomic regions. In one previously uncharacterized associated region, the high-HbF-linked causal variant, rs1010474-C, reduces BACH2 expression and elevates HbF levels. Direct perturbation or inhibition of BACH2 likewise increases HbF expression. Mechanistically, BACH2 restrains activation of the HbF-encoding γ-globin genes, while loss of BACH2 enhances NRF2 chromatin occupancy and promotes the formation of activation foci at the γ-globin genes. Although BACH2 and NRF2 binding motifs in the γ-globin promoters overlap, they can be selectively edited to activate or repress γ-globin, respectively, and do so independently of BCL11A. These findings illustrate how human genetic variation continues to advance our understanding of therapeutically-relevant regulatory mechanisms underlying HbF expression.

## Introduction

During human development, haemoglobin transitions shortly after birth from the fetal form (HbF; containing γ-globin, a β-like globin encoded by *HBG1* and *HBG2*) to the adult form (HbA; containing β-globin encoded by the *HBB* gene). This process is referred to as the fetal-to-adult haemoglobin switch^1^. Persistently increased production of HbF after infancy can ameliorate clinical symptoms in common and life-threatening disorders arising from *HBB* mutations, including sickle cell disease and β-thalassaemia. While the ameliorating effects of HbF in these haemoglobin disorders have been known for decades, the mechanisms regulating HbF and strategies to target them have only recently been elucidated. Initial genome-wide association studies (GWASs) revealed three regions associated with HbF levels— the *HBB* locus on chromosome 11, the *HBS1L-MYB* locus on chromosome 6 and the *BCL11A* locus on chromosome 2^10–12^. These studies prompted functional follow-up that revealed BCL11A as a key and direct repressor of *HBG1/2* transcription^2^. Subsequently, additional insights have emerged from the study of rare loss-of-function mutations impacting BCL11A^13,14^; transcriptional regulatory elements necessary for erythroid expression of BCL11A^15,16^; upstream regulators of BCL11A expression^17–19^; and analysis of the mechanisms by which BCL11A alters transcription^20–22^. Targeting of BCL11A, its erythroid-specific enhancer or its binding sites in the *HBG1/2* promoters using genome editing or gene therapy approaches have emerged as effective treatment strategies for sickle cell disease and β-thalassaemia^3–8^. Moreover, functional studies in cellular models of other HbF regulators have provided insights^9^, such as the identification of ZBTB7A as another direct repressor of the γ-globin genes^22,23^.

Despite substantial progress in understanding HbF regulation, largely derived from studies in cell lines and mouse models, many fundamental questions about how HbF is regulated in humans *in vivo* remain unanswered. The majority of genetically validated regulators of HbF expression that have been identified to date are repressors. However, the precise mechanisms by which the γ-globin genes get activated remain incompletely understood. Despite the progress in understanding the mechanisms underlying HbF expression, many *bona fide* regulators of this process likely remain undefined.

To address these limitations, we conducted a multi-ancestry GWAS of HbF levels in 28,279 individuals from diverse populations. We identified BACH2 as a genetically nominated regulator of HbF expression that acts by restraining NRF2-mediated transcriptional activation of the γ-globin genes. Collectively, these findings define a role for the BACH2–NRF2 axis in regulating HbF and suggest potential therapeutic avenues for the haemoglobin disorders.

## Results

### Multi-ancestry GWAS of HbF

To gain insights into the regulation of HbF expression, we conducted a large-scale multi-ancestry GWAS meta-analysis, which included 28,279 individuals from 11 distinct cohorts spanning European (EUR, n = 22,882, 7 cohorts), African (AFR, n = 4,005, 3 cohorts) and Asian (Thai, n = 1,392, 1 cohort) ancestries (**Fig. 1a and Supplementary Table 1**). In the GWAS meta-analysis, we observed no inflation in test statistics (genomic inflation factor (λ) = 0.99, linkage disequilibrium (LD) score regression intercept = 0.98; **Extended Data Fig. 1a– g**). We identified 91 conditionally independent signals associated with HbF levels across 12 windows, each defined as 1 Mb genomic intervals centred on the corresponding lead variants (eight windows at the genome-wide significance threshold, P < 5 × 10^-8^, and four windows at the suggestive threshold, P < 1 × 10^-6^) (**Fig. 1b and Supplementary Tables 2– 4**). We annotated these windows with genes that satisfied a combination of criteria: (1) distance from a significantly associated variant; (2) long-range interaction data linking regulatory elements to genes in erythroid cells by promoter capture Hi-C (PCHi-C); (3) correlations between gene expression and chromatin accessibility (RNA-sequencing (RNA-seq) and assay for transposase-accessible chromatin with high-throughput sequencing (ATAC–seq) correlations) in haematopoietic cells; and (4) expression quantitative trait loci (eQTLs) from whole blood (**Supplementary Table 5**). Beyond established HbF loci (*HBB, HBS1L-MYB* and *BCL11A*), we identified loci near known HbF regulators that had not been previously reported in GWAS, including *ZBTB7A* and *KLF1*, and loci carrying genes that have not previously been implicated in HbF regulation, including *CTC1, USP34* and *BACH2*.

**Fig. 1:**
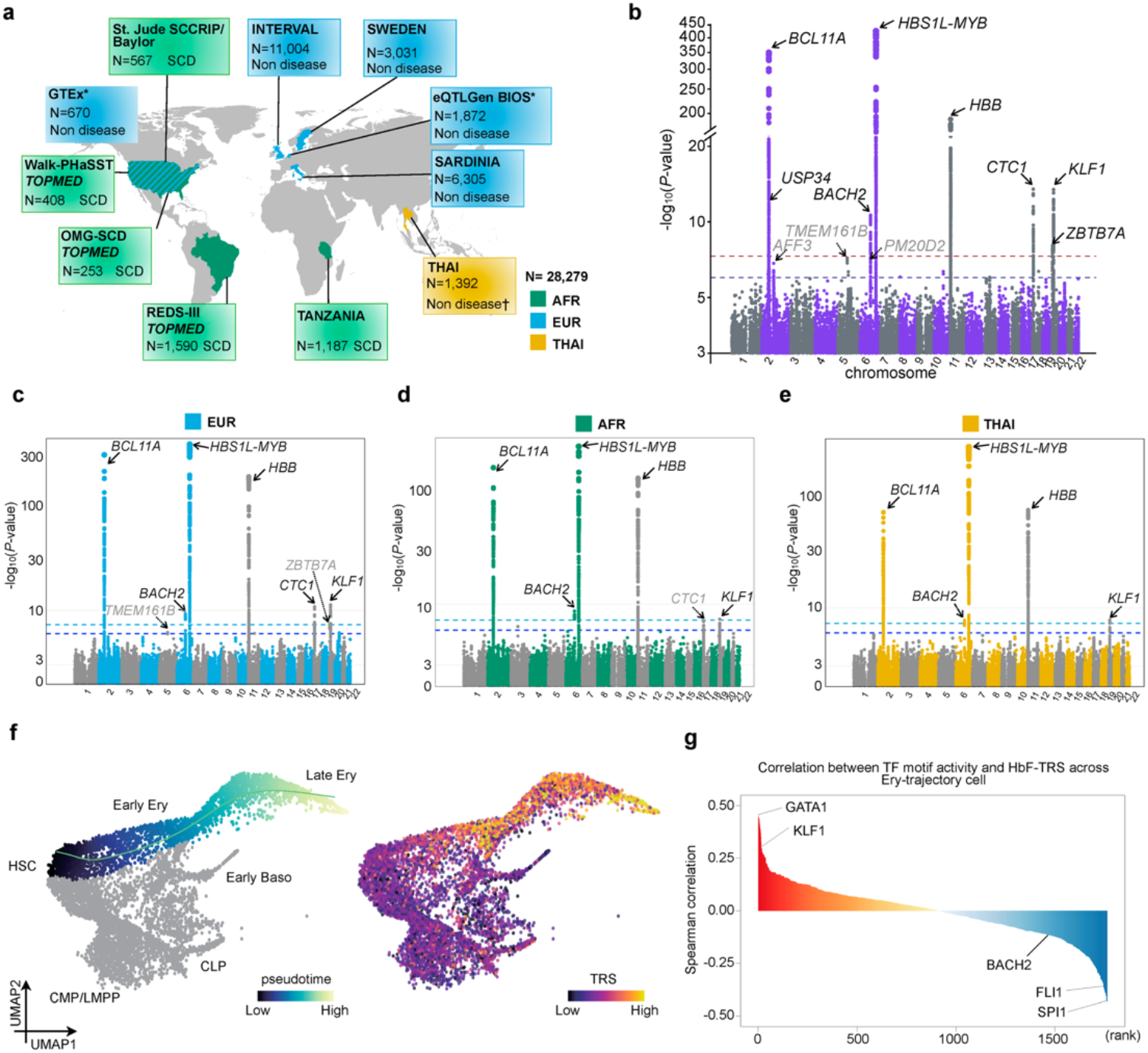
GWAS of HbF across global populations with enrichment of fine-mapped variants in erythroid cells. **a**, Population geography of included studies and associated sample numbers. Note that the Thai population included a proportion of individuals selected for elevated HbF; where indicated by an asterisk, select cohorts were included from the eQTLGen consortium (***Methods***). n = 28,279 individuals. **b**, A fixed-effect meta-analysis of HbF identified several unexplored loci. The gene symbols shown are the most likely impacted gene nominated using several approaches. *P*-values are derived from two-sided sample-size-weighted Z tests implemented in METAL. Genome-wide significant loci (*P* < 5 × 10^-8^) are labeled in black with solid arrows, and suggestive loci (*P* < 1 × 10^-6^) are labeled in gray with dashed arrows. Dashed lines are drawn at *P* = 5 × 10^-8^ and *P* = 1 × 10^-6^ to indicate genome-wide significant and suggestive cutoffs, respectively, which are Bonferroni corrected by a factor of 10^6^. **c-e**, Ancestry-specific analyses using MAMA (Methods) for the European (**c**; EUR), African (**d**; AFR) and Thai (**e**) ancestry backgrounds, respectively. *P*-values are computed from the standard two-sided Wald tests for the effect estimates from MAMA’s multi-ancestry shrinkage estimator. **f**, SCAVENGE analysis using single-cell ATAC–seq (scATAC–seq) data^54^ of haematopoietic differentiation, showing strong enrichment of the TRS in the mid-late erythroid population. HSC, hematopoietic stem cell; LMPP, lymphoid-primed multipotent progenitor; CMP, common myeloid progenitor; baso., basophils; ery., erythroid cells. **g**, Spearman correlations between SCAVENGE TRS and chromVAR TF motif enrichment scores across erythroid trajectory cells (y axis) ranked in order of highest to lowest to identify co-regulated TF motifs.

We next conducted ancestry-specific analyses (**Supplementary Table 6**) using the multi-ancestry meta-analysis (MAMA) approach^24^ for EUR (**Fig. 1c**) or AFR (**Fig. 1d**) ancestry, as well as for the Thai cohort (**Fig. 1e**). We observed largely conserved association signals at the major loci, including at the *BCL11A, HBS1L-MYB, HBB, BACH2* and *KLF1* loci (**Supplementary Table 6**). Among these loci with consistently significant MAMA *P* values across all ancestries, *BACH2* is the only locus not previously implicated in HbF regulation.

We estimated the heritability of HbF levels using LD-adjusted kinship (LDAK), a framework that models per-variant contributions by accounting for both allele frequency and local LD structure. It estimated heritability to be 0.19 (s.e. = 0.058) across all cohorts. For specific ancestry groups, the EUR cohorts show a heritability of 0.25 (s.e. = 0.047), 0.23 for AFR (s.e. = 0.26) and 0.40 for the Thai population (s.e. = 0.32). The higher trait heritability in the Thai population might be attributable to the enrichment for individuals with high HbF levels, which might bias the heritability estimate. After analysis of heritability enrichments for different histone modifications or genomic regions, the major enrichments were seen in putative enhancer elements, suggesting that single-nucleotide polymorphisms (SNPs) residing in and potentially altering regulatory elements contribute most to the currently observed heritability in HbF levels (**Extended Data Fig. 2a**).

Given the well-powered insights from genetic analysis of blood cell phenotypes across populations^25,26^, we examined the extent to which genetic variation associated with HbF levels might also have genetic overlap with these phenotypes (**Extended Data Fig. 2b** and **Supplementary Table 7**). We observed positive genetic correlations between HbF levels and multiple white blood cell phenotypes, including white blood cell (*P* = 0.007) basophil (*P* = 0.015), lymphocyte (*P* = 0.019), monocyte (*P* = 0.026) and neutrophil (*P* = 0.007) counts. A positive correlation trend was also observed for red blood cell count, although this did not reach statistical significance.

### Cellular contexts for HbF variants

We next sought to gain insights into the cellular contexts underlying HbF-associated variation. We fine-mapped variants in each 1 Mb window centred on conditionally independent variants to identify credible sets of potential causal variants. Half of the locus windows had 95% credible sets that were of high resolution with 10 or fewer variants (**Extended Data Fig. 2c, Supplementary Table 8**). These credible set variants are primarily localized to introns and intergenic regions (**Extended Data Fig. 2d**). We then used the SCAVENGE^27^ approach to identify relevant cell states in which the fine-mapped variants showed significant co-localization with accessible chromatin across human haematopoiesis. SCAVENGE integrates fine-mapping posterior inclusion probabilities and singlecell chromatin states, computing the trait relevance score (TRS)—a scaled and normalized metric to quantify the closeness of a cell, over a cell similarity network, to those with the most enrichment of high-confidence variants (that is, trait relevance). We identified a strong enrichment in erythroid cells compared with in other haematopoietic cell types (**Extended Data Fig. 2e-f**). At the single-cell resolution, using a pseudotime projection of human erythropoiesis, we found a strong enrichment at the mid-maturation stages of erythropoiesis (**Fig. 1f**). Given these enrichments, we sought to define co-regulated transcription factor (TF) motifs. Spearman correlations between the SCAVENGE TRS and chromVAR^28^ TF motif enrichment scores across erythroid cells were calculated **(Fig. 1g, Supplementary Table 9**). Notably, KLF1 and GATA1 motifs were highlighted and both TFs are critical in HbF regulation^29^. Collectively, these results highlight key differentiation stages and transcriptional regulatory networks involved in HbF-associated genetic variation.

### High-HbF variant rs1010474-C reduces BACH2 expression

To prioritize loci for further analysis, we integrated evidence from the GWAS meta-analysis and ancestry-specific association analyses. Eight genome-wide significant loci were identified across the meta-GWAS, five of which were also significant in ancestry-specific analyses using MAMA, supporting their robustness across populations. Four of these loci (*BCL11A, HBS1L-MYB, HBB*, and *KLF1*) contain established HbF regulators, whereas *BACH2* was the only previously unreported locus consistently associated with HbF levels across all ancestry-specific cohorts (**Extended Data Fig. 3a**). We therefore prioritized *BACH2* for further investigation. To refine the association signal and identify causal variants, we performed multi-ancestry fine-mapping using MultiSuSiE^30^, a multi-ancestry extension of the SuSiE fine-mapping framework^31^. Analysis of the 1 Mb genomic region centred on *BACH2* identified a credible set comprising 32 tightly linked variants (**Extended Data Fig. 3b**). Owing to extensive LD in this region, statistical fine-mapping alone could not define a likely causal variant or small set of variants, so we sought to integrate these results with functional genomic annotations to further prioritize likely causal variants at this locus. To this end, we intersected the fine-mapped variants across all ancestries based on chromatin accessibility (ATAC–seq) profiles from primary human haematopoietic cells. Among the 32 credible set variants, rs1010473 and rs1010474 have relatively high posterior inclusion probabilities and are the only two that consistently co-localize with accessible chromatin in multiple haematopoietic cell types (**Extended Data Fig. 3c**). To assess the regulatory effects of these naturally occurring variants, we generated a lentiviral reporter containing the putative causal variants in this regulatory element upstream of a minimal promoter. We demonstrated that this regulatory element functions as an enhancer during human erythroid differentiation and the high-HbF-associated allele (C) at rs1010474 reduces enhancer activity (**Fig. 2b and Extended Data Fig. 3d**).

**Fig. 2:**
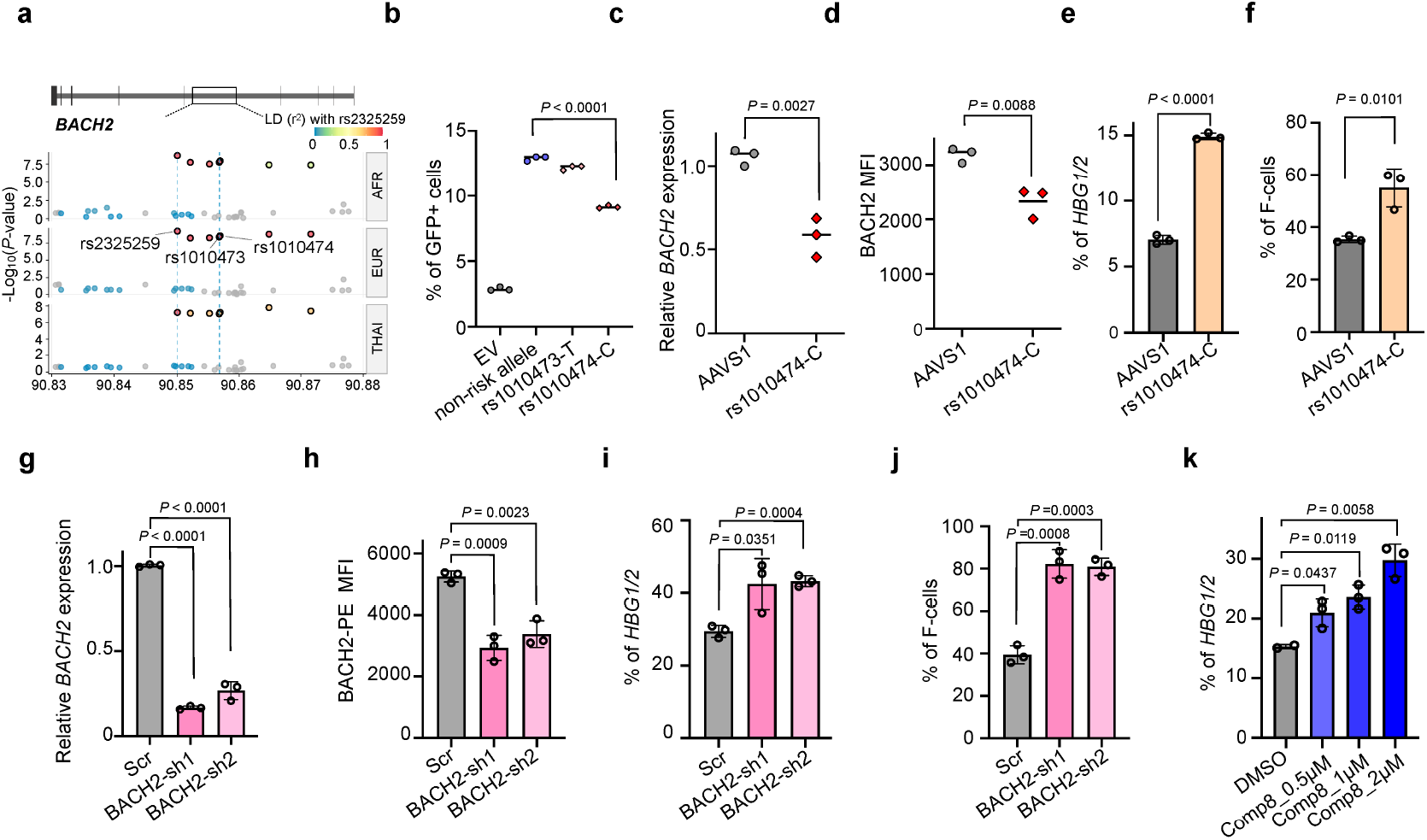
Dissection of BACH2 as a regulator of HbF. **a**, Magnified regional association plots of multi-ancestry fine-mapping at the BACH2 locus. MAMA association signals (−log10*P*-value) are shown for the AFR (n = 4,005), EUR (n = 22,882) and Thai (n = 1,392) cohorts. Multi-SuSiE-prioritized candidate causal variants are indicated by black-bordered circles and coloured by LD (r) with rs2325259, as shown by the colour scale; rs2325259, rs1010473 and rs1010474 are further highlighted by vertical blue lines. **b**, lentiviral reporter assay showing reduced enhancer activity of BACH2 putative causal variants, measured as the percentage of GFP-positive cells on day 6 of erythroid differentiation. n = 3 independent experiments. **c-d**, Relative *BACH2* mRNA (**c**) and intracellular BACH2 protein (**d**) levels in human primary CD34^+^ HSPCs three days after adenine base editing of high-HbF-linked rs1010474-C, compared to *AAVS1* control. MFI, mean fluorescence intensity n = 3 independent experiments. **e-f**, *HBG1/2* expression proportion (**e**) and F-cell frequency (**f**) on day 13 of erythroid differentiation in AAVS1-and rs1010474-C-edited HSPCs. The HBG1/2 expression proportion was calculated relative to total HBG1/2 + HBB expression here and throughout this study. n = 3 independent experiments. **g-h**, Relative *BACH2* mRNA (**g**) and intracellular BACH2 protein (**h**) levels in human primary CD34^+^ HSPCs 4 days after transduction with two BACH2 shRNAs or scramble control shRNA (Scr). MFI, mean fluorescence intensity. n = 3 independent experiments. **i-j**, *HBG1/2* expression proportion (**i**) and F-cell frequency (**j**) on day 13 of erythroid differentiation in HSPCs transduced with BACH2 shRNA and scrambled control shRNA. n = 3 independent experiments. **k**, Proportion of *HBG1/2* expression on day 13 of erythroid differentiation following treatment with the BACH1/2 inhibitor Compound 8 (Comp8). n = 2 for DMSO, n= 3 for comp8 treatment, independent experiments. All data are presented as mean ± standard deviation (SD). Differences between groups were assessed using unpaired two-tailed t-tests.

To define the role of this regulatory element, we performed CRISPR–Cas9-mediated deletion using two pairs of guide RNAs (ENH del1 and ENH del2; **Extended Data Fig. 3e,f**). In the bulk-edited cell population, approximately 40% of BACH2 alleles carried the intended deletion, resulting in reduced BACH2 mRNA and protein levels (**Extended Data Fig. 3g–i**), as well as an induction of HbF, without any disruption of erythroid differentiation (**Extended Data Fig. 3j–l**). Notably, no other genes in the topologically associating domain containing this regulatory element were impacted (**Extended Data Fig. 3m**). To examine the impact of allelic variation at this regulatory element, we used an adenine base editor (ABE8e) to introduce a T>C conversion at rs1010474 with around 40% bulk editing efficiency (**Extended Data Fig. 3n**). We observed reduced levels of BACH2 mRNA and protein after editing (**Fig. 2c,d**), accompanied by induction of HbF (**Fig. 2e,f**), and without any obvious perturbation of erythropoiesis (**Extended Data Fig. 3o**). These findings demonstrate that the rs1010474-C is probably the causal functional variant at this locus, which acts by reducing BACH2 expression within erythroid progenitors, thereby increasing HbF.

### BACH2 prevents HbF activation

Having identified that reduced expression of BACH2 appears to be the key functional target of the high-HbF-associated variant rs1010474-C, we sought to define the mechanisms by which BACH2 regulates the globin genes. Although the related TF BACH1 has been implicated in globin gene regulation^32^, previous work assessing the role of BACH2 in this process is lacking. We initially used short hairpin RNA (shRNA) knockdown to reduce *BACH2* mRNA by up to 80% and protein by up to 50% in primary human hematopoietic stem and progenitor cells (HSPCs) and throughout erythroid differentiation (**Fig. 2g,h and Extended Data Fig. 4a–c**). Notably, knockdown of *BACH2* resulted in minor alterations in erythroid differentiation, but induced HbF during erythroid differentiation, as assessed by both *HBG1/2* mRNA and HbF-containing erythroid cells (F cells; **Fig. 2i,j** and **Extended Data Fig. 4d–h**). To further support these observations, we treated differentiating erythroid cultures with compound 8^33^, a BACH1/2 inhibitor that antagonizes BACH-mediated transcriptional repression of NRF2 target genes. While treatment of erythroid cells at different doses had little or no impact on erythroid differentiation (**Extended Data Fig. 4i,j**), we observed a dose-dependent induction of HbF (**Fig. 2k and Extended Data Fig. 4k**), further reinforcing the role of BACH2 in HbF regulation.

Finally, we sought to reciprocally increase BACH2 expression and assess its role in globin gene expression. To compare cells with different levels of BACH2 expression, we sorted the top (BACH2-GFP^high^) and bottom (BACH2-GFP^low^) 30% of GFP^+^ transduced cells, where GFP is linked to the *BACH2* cDNA through an internal ribosomal entry site and serves as a proxy for BACH2 expression levels (**Extended Data Fig. 5a,b**). We observed a dose-dependent suppression of HbF levels, measured by mRNA and F cell levels (**Extended Data Fig. 5c,d**). There was some perturbation of erythroid differentiation in this setting, most evident through the high BACH2 (around 400-fold) overexpression (**Extended Data Fig. 5e**). Together, these complementary perturbations establish BACH2 as a direct and dosage-sensitive regulator of HbF.

### BACH2 loss enhances NRF2 occupancy

BACH2 encodes a TF that competes with NFE2-like proteins for binding to small MAF proteins at MAF-recognition elements, thereby modulating gene expression^34^. We prioritized NRF2 and NFE2 among potential NFE2-like proteins, as both have been shown to modulate transcription of the β-globin genes and might therefore alter HbF expression^32,35–38^. To investigate the mechanisms of HbF activation, we mapped BACH2, NRF2 and NFE2 chromatin occupancy using cleavage under targets and release using nuclease (CUT&RUN) analysis in human CD34^+^ HSPCs and differentiating erythroid cells. Genome-wide analysis of chromatin occupancy by these factors revealed that their peaks were broadly distributed across promoter, distal intergenic and intronic regions (**Extended Data Fig. 6a**). Within the β-globin loci, BACH2 occupied the γ-globin promoter and its intronic regions before differentiation, suggesting that it restricts γ-globin activation through these cis-regulatory elements. After erythroid differentiation, BACH2 binding increased at the γ-globin promoter and extended to multiple DNase-I-hypersensitive sites within the locus control region (LCR) (**Fig. 3a,b and Extended Data Fig. 6b**), suggesting that it regulates γ-globin through long-range chromatin interactions across the β-globin locus. NRF2 similarly occupied the γ-globin promoters and expanded to hypersensitive sites 1–4 within the LCR after erythroid differentiation (**Fig. 3c, d**), showing substantial occupancy overlap with BACH2. NFE2 also occupied the LCR and the γ-globin promoters, with a relatively constant occupancy pattern during differentiation (**Extended Data Fig. 6c-e**). Given the extensive overlap in chromatin binding and the shared dynamic increases during differentiation, we prioritized NRF2 for mechanistic validation.

**Fig. 3:**
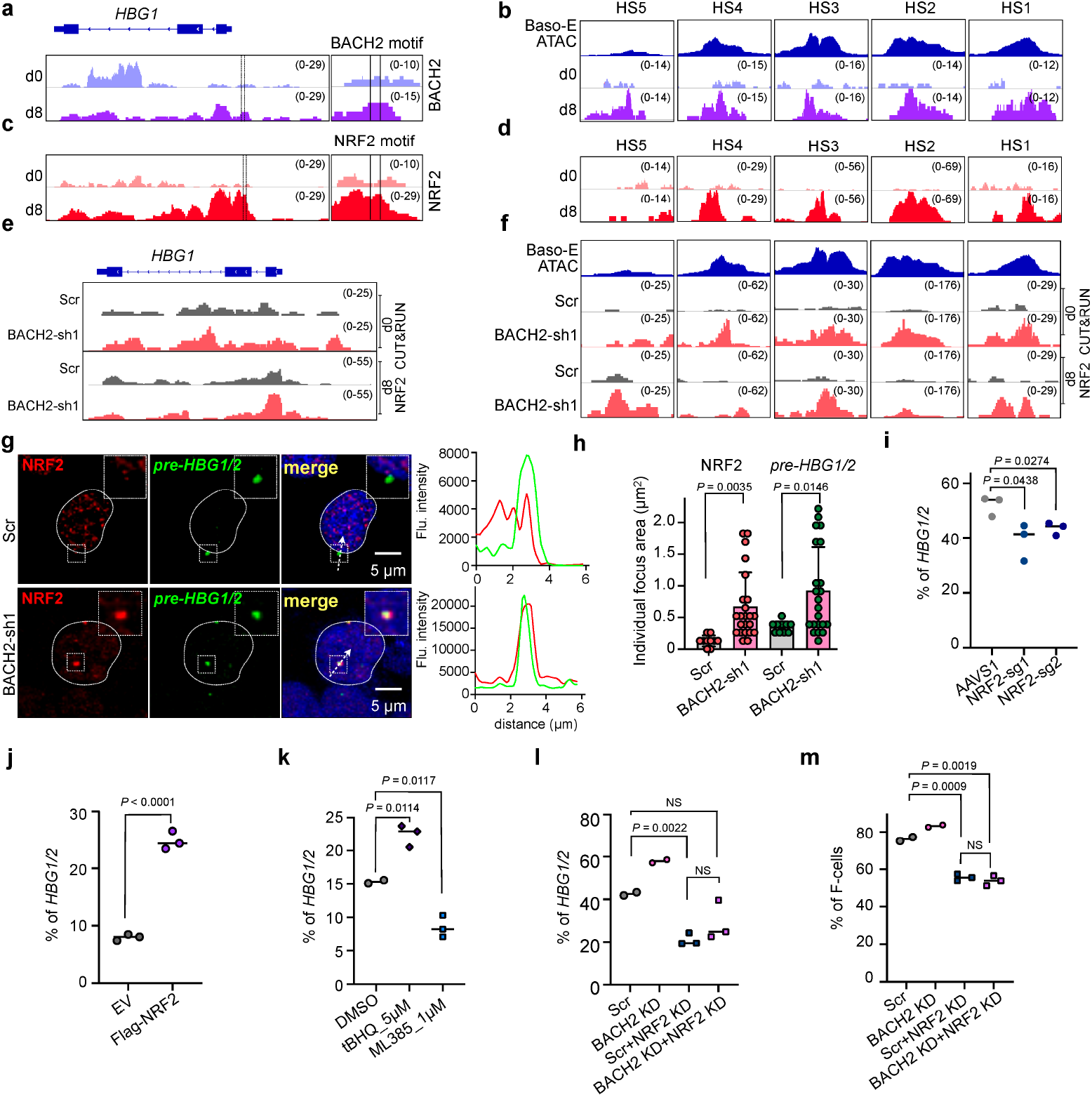
BACH2 loss promotes NRF2 accumulation at the γ-globin locus. **a-b**, BACH2 occupancy at the *HBG1* promoter (a) and β-globin LCR (b) on days 0 and 8 of erythroid differentiation was determined by CUT&RUN.The insets highlight the BACH2 consensus motif at the *HBG1* promoter. Baso-E, basophilic erythroblast. **c-d**, NRF2 occupancy at the *HBG1* promoter (**c**) and β-globin LCR (**d**) on day 0 and 8 of erythroid differentiation. Insets highlight the NRF2 consensus motif at the *HBG1* promoter. **e-f**, NRF2 occupancy at *HBG1* promoter (**e**) and β-globin LCR (**f**) on day 0 and 8 of erythroid differentiation upon BACH2 knockdown. **g**, Co-staining of NRF2 and nascent *HBG1/2* transcripts (*pre-HBG1/2*) in Scr and BACH2 knockdown erythroid cells on day 8 of differentiation. Enlarged views of dashed boxes are shown as upper-right insets. Fluorescence intensity profiles along dashed arrows are shown on the right. **h**, Quantification of NRF2 foci size and *pre-HBG1/2* transcript size in (**g**), Each dot represents one focus. n = 10 cells for control, n = 23 cells for BACH2 knockdown, examined over 3 independent experiments. **i**, *HBG1/2* expression proportion on day 7 of erythroid differentiation in *AAVS1-* and NRF2-edited HSPCs. n = 3 independent experiments. **j**, *HBG1/2* expression proportion on day 11 of erythroid differentiation in empty vector (EV) and NRF2 overexpressed HSPCs. n = 3 independent experiments. **k**, *HBG1/2* expression proportion on day 11 of erythroid differentiation following treatment with the NRF2 activator tBHQ or inhibitor ML385. n = 2 for DMSO, n= 3 for tBHQ or ML385 treatment, independent experiments. **l**, *HBG1/2* expression proportion on day 11 of erythroid differentiation in individual or combined BACH2 and NRF2 knockdown HSPCs. n = 2 for Scr or BACH2 knockdown, n= 3 for combined BACH2 and NRF2 knockdown, independent experiments. **m**, F-cell frequency on day 11 of erythroid differentiation in individual or combined BACH2 and NRF2 knockdown HSPCs. n = 3 independent experiments. All data are presented as mean ± standard deviation (SD). Differences between groups were assessed using unpaired two-tailed t-tests. *P* > 0.05 was considered not significant (NS).

We therefore sought to examine the hypothesis that BACH2 might modulate HbF expression by altering NRF2 binding at the γ-globin locus. We performed NRF2 CUT&RUN analysis after *BACH2* knockdown in HSPCs and differentiating erythroid cells. Loss of BACH2 markedly reduced BACH2 occupancy at the γ-globin promoters in human CD34^+^ HSPCs, with further diminished binding at the γ-globin promoters and multiple hypersensitive sites within the LCR after erythroid differentiation (**Extended Data Fig. 7a,b**). Importantly, loss of BACH2 increased NRF2 occupancy at multiple hypersensitive sites within the LCR and the γ-globin promoters in human CD34 HSPCs, which persisted during erythroid differentiation (**Fig. 3e,f**). NRF2 chromatin immunoprecipitation followed by quantitative PCR (ChIP– qPCR) analysis confirmed enhanced NRF2 occupancy at the γ-globin promoters in differentiated erythroid cells with BACH2 knockdown (**Extended Data Fig. 7c**). These findings indicate that BACH2 normally antagonizes NRF2 binding and its loss enhances NRF2 occupancy at key regulatory elements that might enable the activation of HbF expression.

### BACH2 loss recruits NRF2 to γ-globin

Given the increased NRF2 chromatin occupancy observed after BACH2 loss, we examined whether NRF2 undergoes localized nuclear accumulation at the β-globin locus. NRF2-dependent transcriptional activation is associated with its stabilization and nuclear accumulation^39^. We therefore examined NRF2 subcellular localization by immunofluorescence. Knockdown of *BACH2* led to the appearance of discrete NRF2 nuclear foci, with an average of approximately two foci per cell in human CD34^+^HSPCs (**Extended Data Fig. 7d,e**) and an average of six to seven NRF2 foci per nucleus in K562 cells (**Extended Data Fig. 7f**). We wondered whether these foci correspond to sites of active γ-globin transcription. To examine this, we performed combined NRF2 immunofluorescence and nascent γ-globin (pre-*HBG1/2*) RNA fluorescence in situ hybridization (FISH) analysis in human CD34+ HSPCs and differentiated erythroid cells, and found that the NRF2 nuclear foci colocalize with nascent *HBG1/2* transcripts (**Fig. 3g and Extended Data Fig. 7g**). Quantitative analysis revealed that BACH2-knockdown cells with higher *HBG1/2* expression exhibited larger NRF2 foci and stronger colocalization (**Fig. 3h and Extended Data Fig. 7h**). Notably, analysis of oxidative-stress-related genes after BACH2 knockdown revealed no significant enrichment, and expression of canonical NRF2 targets remained largely unchanged (**Extended Data Fig. 7i,j**). Collectively, these findings support a model in which BACH2 loss allows localized NRF2 recruitment to γ-globin regulatory elements and sites of *HBG1/2* transcription, thereby promoting HbF expression independently of a canonical oxidative stress response.

### NRF2 mediates BACH2 regulation of HbF

As our data suggest a key role of NRF2 in BACH2-mediated HbF regulation, we sought to perturb NRF2 expression and activity in primary human HSPCs undergoing erythroid differentiation (**Extended Data Fig. 8a,b**). CRISPR–Cas9-mediated knockdown of NRF2 reduced HbF levels, including γ-globin (*HBG1/2*) mRNA and F cells (**Fig. 3i and Extended Data Fig. 8c-e**). Reciprocally, increased expression of NRF2 elevated HbF levels (**Fig. 3j and Extended Data Fig. 8f, g**). Consistent with this observation and recent studies^38,40,41^, we found that treatment of cells with the NRF2 inhibitor ML385^42^, a selective NRF2 inhibitor that prevents DNA binding, suppressed HbF expression; by contrast, activation of NRF2 using tBHQ^43^, an NRF2 activator that covalently modifies cysteine residues on KEAP1 to prevent KEAP1-mediated NRF2 degradation—induced HbF expression (**Fig. 3k and Extended Data Fig. 8h-j**). Combined BACH2 and NRF2 depletion did not restore HbF induction (**Fig. 3l,m**), indicating that BACH2 primarily exerts its effects on HbF expression by antagonizing NRF2-mediated HbF activation. Together, these findings demonstrate that BACH2 constrains HbF expression by limiting NRF2-mediated activation of the γ-globin genes.

To further investigate the molecular basis underlying the BACH2–NRF2 regulatory axis, we examined whether the two proteins physically associate. BACH2 was detected in NRF2 cell immunoprecipitants (**Extended Data Fig. 9a,b**), and this association persisted after treatment with DNase I, indicating a DNA-independent interaction (Extended Data **Fig. 9c**). To determine whether this interaction reflects a direct protein–protein interaction, we performed in vitro native polyacrylamide gel electrophoresis (PAGE) analyses using recombinant proteins and found that BACH2 and NRF2 formed a complex (**Extended Data Fig. 9d,e**). Moreover, recombinant protein pull-down assays validated a direct association between BACH2 and NRF2 (**Extended Data Fig. 9f**). Domain mapping revealed that BACH2 associates with NRF2 through its bZip domain (**Extended Data Fig. 9g**). Supporting the specificity of this interaction, no association was detected between BACH2 and NFE2 (**Extended Data Fig. 9h,i**). Together, these findings support a model in which BACH2 constrains NRF2 activity within a common regulatory network, and loss of BACH2 enables NRF2-mediated activation of γ-globin (*HBG1/2*) transcription.

### Editing BACH2/NRF2 motifs modulates HbF

On the basis of our chromatin occupancy data and previously reported BACH2- and NRF2-binding motifs, we defined overlapping but functionally distinct binding sites for these factors at the γ-globin promoters. Specifically, BACH2 has a consensus motif centred around the −94 to −106 region, and NRF2 has a motif centred around the −96 to −105 region (**Fig. 4a**). To investigate their function roles, we edited the potential binding sites within the −94 to −106 region using either adenosine base editors (ABE) or cytosine base editors (CBE), followed by assessment of HbF expression (**Extended Data Fig. 10a**). We achieved around 90% –105T>C and >50% –106T>C substitutions using ABE, and around 50% –98G>A, –99G>A and –104G>A substitutions using CBE (**Extended Data Fig. 10b**). Notably, the –105T>C and –106T>C substitutions reduced HbF expression, whereas the –98G>A, –99G>A and – 104G>A substitutions induced HbF expression (**Fig. 4b,c and Extended Data Fig. 10c–e**). Importantly, perturbation of these cis-elements did not impact erythroid differentiation (**Extended Data Fig. 10f**). Altering these potential binding sites exerts differential effects on HbF expression, suggesting that they may differentially modulate the binding preference of BACH2 or NRF2 within this region.

**Fig. 4:**
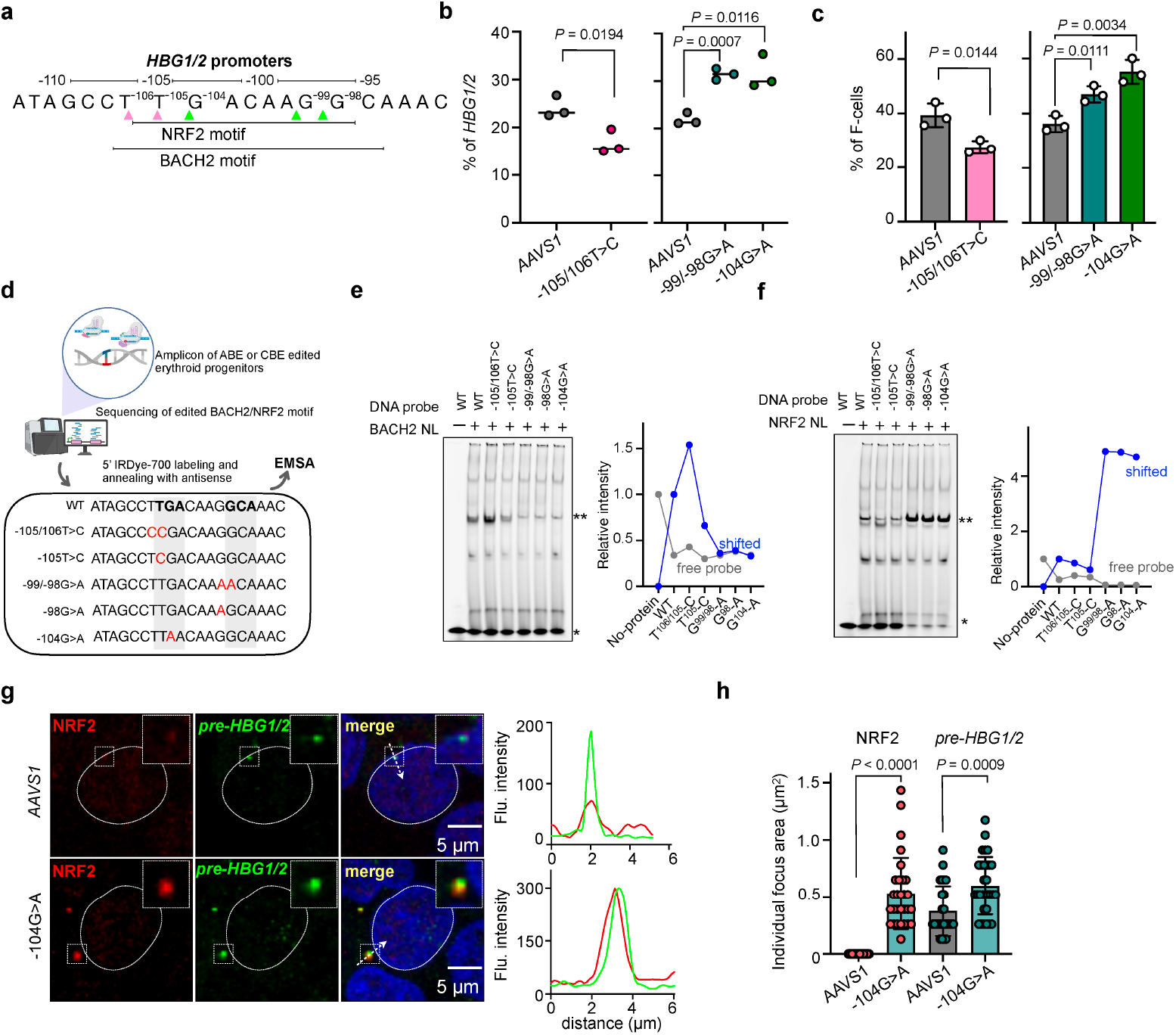
Editing of BACH2/NRF2 motif modulates HbF expression. **a**, Schematic showing the positions of BACH2 and NRF2 motifs in the *HBG1/2* promoters. Green and pink triangles indicate cytosine base editing (CBE) and adenosine base editing (ABE) sites, respectively. Superscript numbers indicate nucleotide positions within the *HBG1* promoter. **b**, Proportion of *HBG1/2* expression on day 11 of erythroid differentiation in *AAVS1* control and BACH2/NRF2 motif-edited HSPCs following ABE and CBE editing. n = 3 independent experiments. **c**, Frequency of F-cells on day 16 of erythroid differentiation in *AAVS1* control and BACH2/NRF2 motif-edited HSPCs. n = 3 independent experiments. **d**, Schematic of EMSA probe design. 20-nt double-stranded probes (WT or edited variants) were generated from sequenced amplicons of base-edited erythroid progenitors. Sequence alignment shows the WT and edited BACH2/NRF2 motifs used for EMSA. **e-f**, EMSA and quantification showing BACH2 (**e**) or NRF2 (**f**) binding to the indicated DNA probes. Double asterisks (**) indicate protein-DNA complexes (shifted); single asterisk (*) indicates free probes. Quantification shows the relative intensity of shifted bands (blue) and free probes (grey). **g**, Co-staining of NRF2 and *pre-HBG1/2* in *AAVS1-* and −104G>A-motif-edited erythroid cells on day 8 of differentiation. Enlarged views of dashed boxes are shown as upper-right insets. Fluorescence intensity profiles along the dashed arrows are shown on the right. **h**, Quantification of NRF2 foci size and *pre-HBG1/2* transcript size in (**g**). Each dot represents one focus. n = 26 cells for *AAVS1*, n = 29 cells for −104G>A-edited group, examined over 3 independent experiments. All data are presented as mean ± standard deviation (SD). Differences between groups were assessed using unpaired two-tailed t-tests. Icons in **d** are created in BioRender. Guo, C. (2026) https://BioRender.com/ggt2bq3.

To assess DNA binding by NRF2 or BACH2, we performed in silico predictions of TF-binding affinities at the edited cis-elements, which revealed that the –105T>C and – 106T>C substitutions were predicted to markedly reduce NRF2 binding, whereas the –98G>A, –99G>A and – 104G>A substitutions were predicted to reduce BACH2 binding (**Extended Data Fig. 10g**). We experimentally validated that this region is capable of binding to both BACH2 and NRF2 using electrophoretic mobility shift assays (EMSAs) (**Extended Data Fig. 10h,i**). To determine whether these variants alter BACH2 or NRF2 recruitment, we performed EMSAs using DNA probes corresponding to the edited sequences (**Fig. 4d**). BACH2 binding was increased by the – 105/–106T>C substitutions but reduced by the –98G>A, –98/– 99G>A and –104G>A substitutions (**Fig. 4e**). By contrast, NRF2 exhibited the opposite binding pattern across the same set of variants (**Fig. 4f**). In parallel, we also observed NRF2 accumulation at the sites of nascent *HBG1/2* transcripts after cytosine base editing (**Fig. 4g,h and Extended Data Fig. 10j**), supporting the role of NRF2 binding in γ-globin activation and HbF induction.

### BACH2 regulates HbF independently of BCL11A

BCL11A is a key repressor of HbF and binds to two motifs within the γ-globin promoters—a distal site (−113 to −118) and a proximal site (−86 to −91)^6,21^. The NRF2 and BACH2 motifs (−94 to −106) are positioned between these two BCL11A sites (**Fig. 5a**). We therefore tested whether BACH2 regulates HbF through BCL11A. We disrupted the +58 erythroid enhancer of BCL11A while simultaneously perturbing BACH2 expression. Knockdown of BACH2 or BCL11A individually promotes HbF expression after erythroid differentiation, whereas simultaneous depletion of both factors resulted in an additive effect, producing higher HbF levels than either single knockdown alone (**Fig. 5b**). To examine whether these proteins physically interact, we performed co-immunoprecipitation assays. Neither BCL11A–BACH2 nor BCL11A–NRF2 interactions were detected (**Fig. 5c,d**). These results indicate that BACH2-mediated regulation of HbF occurs independently of BCL11A and that the two pathways act in parallel (**Fig. 5e**).

**Figure 5:**
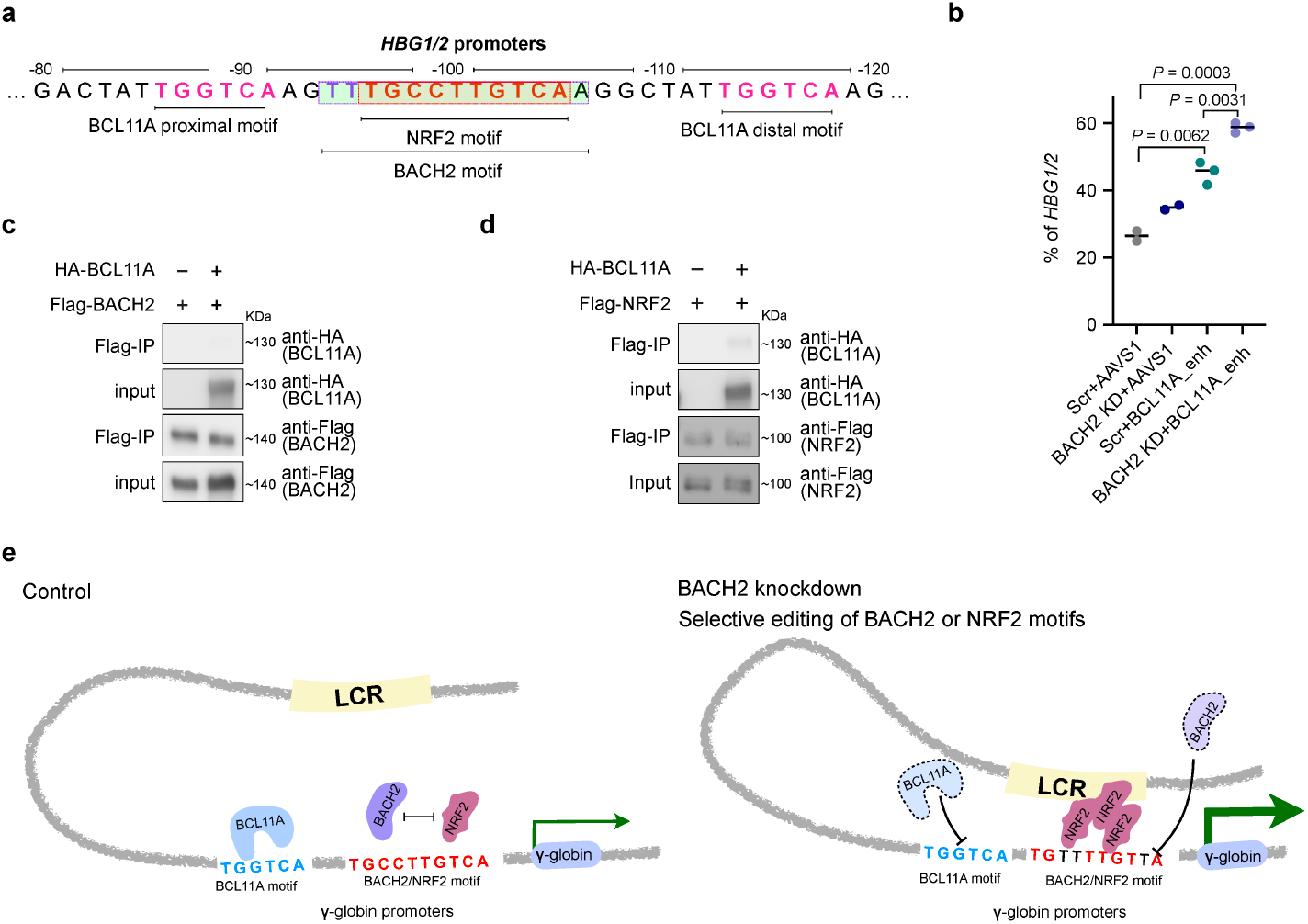
BACH2 regulates HbF independent of BCL11A. **a**, Schematic showing the positions of BACH2 motif, NRF2 motif and BCL11A distal and proximal motif within the *HBG1/2* promoters. **b**, *HBG1/2* expression proportion on day 11 of erythroid differentiation with combined BACH2 and BCL11A knockdown. n = 2 for (scrambled control and BACH2 knockdown) and n = 3 (combined BACH2 knockdown and BCL11A enh disruption) independent experiments. BCL11A enh, disruption of the BCL11A +58 enhancer. **c**, BCL11A does not interact with BACH2. Co-immunoprecipitation was performed using anti-Flag antibody in HEK293 cells overexpressing Flag-tagged BACH2 and HA-tagged BCL11A, followed by immunoblotting with anti-HA antibody. **d**, BCL11A does not interact with NRF2. Co-immunoprecipitation was performed using anti-Flag antibody in HEK293 cells overexpressing Flag-tagged NRF2 and HA-tagged BCL11A, followed by immunoblotting with anti-HA antibody. **e**, Schematic of the working model illustrating the regulation of HbF by BACH2–NRF2 and BCL11A at the γ-globin promoters. Left, BCL11A binds to the distal motifs to repress HbF expression, BACH2 and NRF2 competitively bind to the motif located between the proximal and distal BCL11A motifs to repress or activate HbF expression, respectively. Right, BACH2 knockdown or selectively editing BACH2/NRF2 motifs in γ-globin promoters promotes NRF2 recruitment to HBG1/2 regulatory elements and active transcriptional sites. The arrows indicate activation of HbF, and the blocked arrows indicate repression. All data are mean ± s.d. Differences between groups were assessed using two-sided unpaired t-tests.

## Discussion

Considerable progress in our understanding of HbF regulation has emerged through studies of human genetic variation, enabling therapeutic advances. However, the studies of genetic variation to date have been limited in size to several thousands of individuals at most and have typically been focused on specific ancestry groups^44–46^. Concomitantly, studies of individuals with exceptionally elevated HbF levels have revealed rare genetic variation at the β-globin gene locus and in other genes, including *BCL11A, KLF1* and *ZBTB7A*, which leads to larger increases in HbF levels^13,47–49^. Here, by conducting a large multi-ancestry GWAS meta-analysis of HbF levels, we identify a number of loci underlying variation in HbF levels. Together, these findings further define the genetic architecture of HbF regulation and provide a framework for identifying additional regulatory pathways that may be leveraged for therapeutic HbF induction.

To date, genetic variation in *BACH2* has primarily been associated with autoimmune and allergic diseases^50,51^, with no attention given to its potential roles in HbF regulation. Here, by following up on insights from our human genetic studies, we provide functional evidence that BACH2 restrains NRF2-mediated activation of HbF. Critically, we disentangle the mechanisms through which these factors modulate activation of the γ-globin promoters. These findings establish the BACH2–NRF2 axis as a tractable and potentially therapeutically targetable regulatory node involved in HbF activation. Although NRF2 has been suggested to induce γ-globin expression through the classical oxidative stress response^40^, our findings reveal a distinct mechanism involving the interplay between BACH2 and NRF2, independent of canonical stress-response signalling. Furthermore, we demonstrate that this axis is independent of the well-studied and targeted BCL11A-dependent repressive mechanisms, suggesting synergistic opportunities to modulate HbF expression by targeting both pathways in tandem, such as could be achieved by combining cis-element perturbations for these various regulators in the HBG1/2 promoters, further advancing approaches recently reported from clinical trials targeting the BCL11A binding sites alone^5,6^. While our data support NRF2 as the primary functional partner of BACH2 in this context, we cannot formally exclude contributions from other NFE2-like TFs that might also be involved in γ-globin regulation.

In summary, by studying the genetic basis of variation in HbF levels across diverse populations, we have defined a role for the BACH2–NRF2 axis in enabling HbF activation. While the effect size of the *BACH2* variant on HbF levels is lower in comparison to other loci, such as *BCL11A*, the presence of high-confidence human genetic evidence is a more critical predictor of clinical success than the absolute magnitude of the variant effect size^52^. By providing cross-ancestry genetic evidence and a clear mechanistic framework for BACH2-mediated HbF regulation, our study provides a fundamental basis for further investigation of this pathway and its potential therapeutic relevance. Many further biological mechanisms are likely to be identified through the study of additional loci identified here and through further expansion of such HbF GWAS. Indeed, a recent study has similarly used more-recent genetic association findings to define a distinct regulator of HbF—*ABCC1*^53^. Our study provides an example of how increasingly larger GWAS can enable further insights into even seemingly well-understood and therapeutically important human phenotypes like HbF.

## Data Availability

All data produced in the present study will be released upon publication of this work.

## Acknowledgements

We are grateful to members of the Sankaran laboratory for valuable comments and discussion, as well as S. Orkin and D. Nathan for their inspiration and guidance. We acknowledge early work on these cohorts by J. Verboon, A. Cheng, and C. Fiorini. We thank V. Kuchroo, A. Schnell, and Y. Hou for providing reagents. We thank A. Martin and M. Kanai for their insightful suggestions and helpful discussions on multi-ancestry fine-mapping.

## Funding

Work in the lab of V.G.S. is supported by the Howard Hughes Medical Institute, the New York Stem Cell Foundation, and National Institutes of Health (NIH) grants R01 DK103794, R01 DK101989, R01 HL146500, R01 CA265726, and R01

CA292941, the Gates Foundation, the Edward P. Evans Foundation, Alex’s Lemonade Stand Foundation, Blood Cancer United, and the Leona and Harry Helmsley Charitable Trust. V.G.S. is an investigator of the Howard Hughes Medical Institute.

## Author information

### Contributions

V.G.S., C.-J.G. conceptualized and designed the study. C.-J.G., U.P.A., X.C., L.D.C., B.S.M., S.E., P.D., L.M., R.S., T.S., S.R., P.G.B., D.S.P., E.K., W.J.A., F.A., K.A., A.L.L.P, G.K., Y.Z., S.M.N., V.R.G., M.T.G, M.E.G, A.A.-K, M.J.T., B.C., S.K., C.L.D., E.C.S., P.L., A.B.C.-P., C.M., A.M., A.H.-L., V.A.S., M.J.W., L.F., B.N., A.S.B., V.V., S.N., V.G.S. obtained and provided cohort data. C.-J.G., R.L., H.Y.L. P.L., T.Y., M.A. performed functional studies. U.P.A, X.C., W.X., L.D.C., A.J.L. F.Y., G.A., M.W. performed computational analyses. V.G.S., C.-J.G, U.P.A., X.C., W.X., L.D.C., R.L., H.Y.L. wrote the original draft with input from all authors. V.G.S. provided overall study oversight. All authors were involved in reviewing and editing the manuscript. All authors read and approved the final version of the manuscript.

## Ethics declarations

### Competing interests

During the drafting of the manuscript, D.S.P. became a full-time employee of AstraZeneca. F.A. is an employee and shareholder of Illumina, Inc. M.T.G. has financial interests in Globin Solutions, Inc. and serves as a consultant for Synhale Therapeutics and Third Pole. A.H.L. reports speakers honoraria from Siemens Healthineers and Beckman Diagnostics, as well as participation on an advisory board of Roche Diagnostics, all unrelated to the present work. A.S.B. reports institutional grants from AstraZeneca, Bayer, Biogen, BioMarin, Bioverativ, Novartis, Regeneron and Sanofi. V.G.S. serves as an advisor to Ensoma, Cellarity, and Beam Therapeutics, all unrelated to the present work. The other authors declare no competing interests.

### Data availability

Raw and processed data have been deposited to public databases and will be available at the time of publication. GWAS summary statistics will also be made available at the time of publication.

## Materials and Methods

### Individual GWAS study methods and quality control

Details including study design, genotyping, and imputation methods and quality control for the included studies is in **Supplementary Table 1**. The cohorts relied upon different selection strategies, including unselected individuals from the population (Swedish, SardiNIA^10^, INTERVAL^55^, GTEx^56^, BIOS^57^), individuals with sickle cell disease (Tanzania^45^, Walk-PHaSST^58^, OMG-SCD^59^, REDS-III Brazil^60^, St. Jude Sickle Cell Clinical Research & Intervention Program (SCCRIP)/Baylor^61,62^), or individuals selected from a screened population (Thai). Note that because our meta-analysis includes both general population (European ancestry) and high-HbF (African and Thai ancestry) cohorts, the effect sizes are not comparable across ancestries.

The Thai cohort has a unique study design: from a large general population of ~86,000 individuals, we intentionally screened for high HbF and sampled individuals with HbF levels higher than 2% of total hemoglobin. We then randomly selected other individuals from this same general population to serve as contrast. Individuals with severe forms of thalassemia were excluded. DNA extraction, sequencing, and genotyping were performed after the sample selection. HbF levels for the entire cohort (enriched for individuals with high HbF) were inverse-normalization transformed prior to GWAS analysis.

We examined the relatedness among individuals in TOPMed (including Walk-PHaSST, OMG-SCD, and REDS-III Brazil), Tanzania, and Thai cohorts, and did not find outliers (KING relatedness above 0.0884) to be excluded. Data from the other cohorts do not include individual genotypes, but we have confirmed with their authors that the relatedness has been accounted for in their cohort-specific association studies. Most included samples had HbF measured in the traditional way using high performance liquid chromatography, however two of the included cohorts (BIOS and GTEx) derived the HbF phenotype from expression data (in TPM units) as a ratio of gene expression of (*HBG1* + *HBG2*) / *HBB*. We found this faithfully replicates expected results from the traditionally measured HbF cohorts (see **Supplementary Note**). The included Swedish and Thai populations are previously undescribed cohorts and were analyzed specifically for this study. All participants provided informed consent, and all studies obtained ethical approvals from local ethics review boards. Details about each cohort and their GWAS procedures are described in full in **Supplementary Note**.

All GWAS summary statistics were lifted-over from their respective genome builds to reference genome hg38. Alleles were flipped according to the hg38 build reference allele, and if neither allele was present the variant was removed. Strand ambiguous and non-biallelic SNPs were removed. Minor allele frequency was filtered to >=0.1%. RSIDs were assigned using dbSNP version 144. All models included adjustment for age, sex, and the top 10 principal components derived from the respective cohort. In addition, for the SCD cohort analyses, SCD genotypes and hydroxyurea use were included as categorical covariates in the respective association models.

### Meta-analysis

Fixed effects meta-analysis (FEMA) of all cohorts was performed using METAL r2020-05-05 (github.com/statgen/METAL)^63^ under the sample size-weighted scheme with genomic control, yielding results for 21,849,438 SNPs genome-wide.

For ancestry-specific meta-analysis, we deployed Multi-Ancestry Meta analysis (MAMA, version #16865), which boasts improved power in meta-analysis of different populations with low false positive rates after correcting for cross-population LD and frequency differences^24^. We provided MAMA with reference panels consisting of 1000 genomes^64^ samples (892 AFR and 661 EUR) and Thai whole genome sequenced (WGS) samples (N=198) to compute stratified LD scores (step 1). We then performed two sets of inverse-variance-weighted (IVW) FEMA without genomic control on all European or African cohorts. Step 2 of MAMA was subsequently performed on the summary statistics of the two FEMA and the Thai cohort, along with the LD score matrix from step 1, yielding 3,760,527 intersecting SNPs. Notably, the high correlation of effect estimates observed across populations (**Extended Data Fig. 1e-g**) is in part explicitly induced by MAMA’s underlying model of shared genetic effects. The remaining variability in effect estimates may reflect an interplay of biological and technical factors. These include inherent differences in allele frequencies and LD structures, as well as potential technical mismatches between non-European study cohorts and the reference resources used for imputation and LD estimation.

### Identifying conditionally independent loci and multi-ancestry fine-mapping

Following the official guidance to use the largest available cohort as the LD reference in meta-analytic settings, we created an external LD reference panel from AllOfUs (v5) data^65^ European samples (EUR, N=51,125, downsampled to 10,000) for conditional analysis by GCTA-COJO (v1.94.0)^66^, on the all-cohort FEMA summary statistics with options --cojo-slct --cojo-p 1e-06 --cojo-wind 10000 --cojo-collinear 0.9 --diff-freq 0.6 --maf 0.001. A separate run using an African reference panel (10,000 from AllofUs) was also performed. The conditionally independent loci identified by European-based COJO became our sentinel markers; loci with too much conditional Z-score inflation (>10-fold) were discarded. We examined the 1Mb windows centered on these sentinel markers and labeled them with the nearest or, if known, biologically relevant gene. Once each locus was determined, fine-mapping was performed using FINEMAP^67^ with the parameters: --non-funct --allow-missing --max-num-causal 1. Note that although the use of a European-only reference panel for COJO inevitably introduces LD mismatches for multi-ancestry summary statistics, we only intended for it to coarsely partition independent signals genome-wide, rather than to make precise conditional inference. Rather, downstream variant prioritization was jointly based on MAMA, MultiSuSiE, marginal association patterns, and functional annotations.

Multi-ancestry fine-mapping was performed with MultiSuSiE^30^ on the 1Mb regions centered on *BACH2*. Pre-computed LD matrices from the PanUKB consortium were leveraged for three ancestries: European (EUR), African (AFR), and East Asian (EAS, as a proxy for the Thai cohort). Cohort-specific summary statistics were lifted over to hg19 to match the LD matrices. Within AFR and EUR ancestry, cohorts were meta-analyzed via the inverse variance-weighted scheme by METAL without genomic control to meet MultiSuSiE’s assumptions. Three- and two-ancestry (EUR and AFR) finemapping were then performed assuming component number L {3, 5, 10, 12, 15} with custom scripts.

### Heritability and genetic correlation analyses

LD scores were established from AllofUs (v5) LD panels as described above using maximum 1cM window positions. LDSC^68^ was performed on summary statistics restricted to high quality, HapMap 3 variants. LDAK^69^, was used to estimate heritability using the thin and BLD models appropriate for ancestry. Genetic correlations with blood cell traits were estimated using LDAK, using the thin model. Summary statistics for blood cell traits were obtained from the published BCX2 consortium summary statistics available at http://www.mhi-humangenetics.org/en/resources/.

### SCAVENGE

SCAVENGE^27^ was performed (https://sankaranlab.github.io/SCAVENGE/articles/SCAVENGE) using custom scripts. We first obtained fine-mapped variants with posterior probabilities of causality from FINEMAP. We then collected single-cell ATAC-seq data from 10 individuals, comprising 33,819 cells across 23 annotated cell populations^54^. Slingshot^70^ was used to reconstruct the developmental progression of erythroid lineage and to assign pseudotime to each cell along the trajectory. Using the fine-mapped variants and single-cell chromatin accessibility data as input, we calculated trait relevance scores (TRS) for individual cells. In parallel, we generated pseudo-bulk profiles based on cell-type annotations to construct pseudo-bulk ATAC-seq datasets as previously described^71^, and used these data as input for peak calling with MACS2^72^.

### Promoter Capture Hi-C (PCHi-C)

For annotating genomic windows with genes, we used a published promoter capture Hi-C (PCHi-C)^73^ dataset spanning 15 haematopoietic cell types. We retained chromatin looping interactions with a CHiCAGO score > 5.

### Human primary HSPC culture and in vitro erythroid differentiation

Human CD34^+^ HSPCs from mobilized peripheral blood of healthy adults were obtained from the Cooperative Center of Excellence in Hematology at the Fred Hutchinson Cancer Research Center. The primary HSPCs were thawed into a maintenance medium consisting of a StemSpan II base (StemCell Technologies), CC100 (StemCell Technologies), 50 ng/mL human TPO (Pepro Tech) and 1% penicillin/streptomycin (Life Technologies) and 1% of L-Glutamine (Life Technologies)^74,75^.

After the maintenance phase, primary human HSPCs were differentiated using the three-phase culture system previously described^76,77^. First, a base erythroid medium was created by supplementing IMDM (Gibco) with 2% human AB plasma (SeraCare), 3% human AB serum (Life Technologies), 3 U/mL heparin, 10 µg/mL insulin, 200 µg/mL holo-transferrin, and 1% penicillin/streptomycin. From days 1-7 in erythroid media, this base medium was further supplemented with 3 U/mL EPO, 10 ng/mL human SCF, and 1 ng/mL IL-3. From days 7-12, this base medium was further supplemented with 3 U/mL EPO and 10 ng/mL human SCF. After day 12, the base medium was supplemented with 1 mg/mL total of holo-transferrin and 3 U/mL of EPO.

### Cell line culture

HEK293T cells were cultured in Dulbecco’s Modified Eagle’s Medium (DMEM) supplemented with 10% FBS and 1% penicillin/streptomycin. K562 cells were cultured in Iscove’s Modified Dulbecco’s Medium (IMDM) supplemented with 10% FBS and 1% Penicillin-Streptomycin.

### Genome editing of human primary HSPCs

Genome editing was performed in human primary CD34^+^ HSPCs using the 4D-Nucleofector system (Lonza) with the P3 Primary Cell 4D-Nucleofector X Kit S (Lonza). For CRISPR-Cas9-mediated enhancer deletion, ribonucleoprotein (RNP) complexes were assembled by combining 100 pmol Cas9 protein (IDT) with 100 pmol chemically modified sgRNAs (Synthego) and incubating at room temperature for 15 min. RNPs were delivered into CD34^+^ HSPCs with the P3 nucleofection reagent supplemented with nucleofection enhancer (IDT) at a 20:1 ratio, using program DZ-100 on the 4D-Nucleofector system. To generate the high-HbF–associated variant rs1010474, 20 µg of adenosine base editor (ABE8e)^78^ protein was incubated with 100 pmol chemically modified sgRNA (Synthego) at room temperature for 20 min. The assembled ABE-RNP complexes were delivered into CD34^+^ HSPCs using P3 reagent and program DZ-100. For editing NRF2/BACH2 motifs within the *HBG1/2* promoters, 2 µg of *in vitro* transcribed ABE8e or TadCBEb^79^ were mixed with 100 pmol chemically modified sgRNAs (Synthego) and nucleofected into CD34^+^ HSPCs using program DS-130. Electroporated cells were harvested 3 days after nucleofection for genomic DNA and RNA extraction using the AllPrep DNA/RNA Micro Kit (Qiagen) following the manufacturer’s instructions. All guide RNA sequences used for genome editing are listed in **Supplementary Table 11**.

### BACH2 shRNA knockdown and overexpression cloning and lentivirus packing

To knock down BACH2, DNA sequences for shRNAs were designed by the GPP Web Portal online tool (https://portals.broadinstitute.org/gpp/public/). DNA oligos of shRNA sequences for targeted RNAs or scramble shRNAs were individually cloned into pLKO.1-GFP vector. For overexpression, the human *BACH2* coding sequence was synthesized by Azenta and inserted into the HMD lentiviral vector. All the plasmids were validated by whole plasmid sequencing by Plasmidsaurus. Lentiviral particles were produced as previously described. Briefly, 5 × 10^6^ HEK293T cells in a 10 cm dish were co-transfected with 10 µg pLKO.1 shRNA constructs or overexpression construct, 7.5 µg of psPAX2 and 3 µg pMD2.G plasmids. The supernatant containing viral particles was harvested twice at 48h and 72h after transfection, then filtered through Millex-GP Filter Unit (0.45 µm pore size, Millipore) and concentrated by ultracentrifugation (24,000 rpm, 2 h, 4 °C). Concentrated virus was used to transduce HSPCs in the presence of 8 µg/mL polybrene (Millipore) by spinfection (2,000 rpm, 1.5 h, RT). Transduced cells were sorted based on GFP expression by fluorescence activated cell sorting (FACS) and subsequent to erythroid differentiation and functional analyses.

### RNA isolation and RT-qPCR

RNA was collected from cultured cells using the Total RNA Purification Micro Kit (Norgen Biotek) with DNase I treatment according to the manufacturer’s protocol. The cDNA synthesis was carried out using PrimeScript RT Master Mix (TaKaRa) according to the manufacturer’s protocol. RT-qPCR was run on the CFX96 or CFX384 Real Time System (BioRad) using iQ SYBR Green Supermix (BioRad) following kit instructions. The relative expression of different sets of genes was quantified to *ACTB* mRNA, with control samples serving as the reference. Primer pairs for RT-qPCR are listed by gene in **Supplementary Table 11**.

### Cell staining for Flow cytometry analysis

To assess erythroid differentiation, transduced or genome-edited HSPCs were collected at the indicated time points. Following a DPBS wash, cells were stained on ice for 30 min in FACS buffer (DPBS + 0.1% BSA) with anti-CD71_BV421 (BioLegend) and anti-CD235a_APC-Cy7 (BioLegend). Cells were then washed and resuspended in FACS buffer for flow cytometry analysis. For BACH2 intracellular staining, 3 × 10^5^ human primary HSPCs were collected and washed with DPBS, then fixed in 4% paraformaldehyde (PFA) in PBS for 15 min. Cells were permeabilized with 0.2% Tween-20 in PBS for 10 min and stained with BACH2-PE (BioLegend) in 0.2% Tween-20 in PBS for 1 h at room temperature. Cells were subsequently washed and resuspended in FACS buffer for flow cytometry analysis. To quantify F-cell frequency in transduced or genome edited HSPCs undergoing erythroid differentiation, cells were fixed in 4% paraformaldehyde (PFA) in PBS for 15 min, permeabilized with 0.2% Tween-20 in PBS for 10 min, and stained with anti-HbF-PE (BD Biosciences) or anti-HbF-APC (invitrogen) in PBS containing 0.1% BSA for 1 h at room temperature. Cells were subsequently washed and resuspended in FACS buffer for flow cytometry analysis. For each sample, 30,000-50,000 events were acquired on a BD LSRFortessa (BD Biosciences), and analyzed using FlowJo software (v10.8.1, BD Biosciences).

### Lentiviral reporter assays

To investigate the functional impact of *BACH2* regulatory variants, lentiviral reporter constructs were generated by cloning the *BACH2* regulatory element containing either the non-risk allele or the risk alleles upstream of a minimal TATA-box promoter (miniP) driving eGFP expression as a functional readout. Lentiviral particles were produced via transient transfection in HEK293T cells and concentrated by ultracentrifugation. To ensure experimental consistency, normalization was performed at the genomic level by quantifying the number of integrated proviruses. Specifically, genomic DNA (gDNA) was extracted from transduced K562 cells and the proviral copy number was determined by qPCR using the Lenti-X™ Provirus Quantitation Kit (Takara) utilizing a standard curve of CT values versus copy number for precise titration. Subsequently, titer-normalized lentiviruses were used to transduce primary HSPCs which then underwent *in vitro* erythroid differentiation. The regulatory activity of the tested elements was quantitatively assessed by measuring GFP expression via flow cytometry throughout the differentiation.

### Co-immunoprecipitation and western blotting

HEK293T or K562 cells (2 × 10^7^) expressing Flag-NRF2, HA-BACH2, or their truncation constructs were harvested and lysed in 1 mL lysis buffer (50 mM Tris pH 7.4, 150 mM NaCl, 0.05% Igepal, 0.5% NP-40, 0.5 mM PMSF, and protease inhibitor cocktail), followed by two 10-s pulses of sonication. Lysates were pre-cleared with 15 µL protein G beads (Invitrogen) for 30 min at 4 °C. Pre-cleared lysates were incubated with 5 µg anti-Flag M2 antibody (Sigma) or 5 µg isotype IgG control (Santa Cruz) for 3-4 h at 4 °C, and then incubated with protein G beads for an additional 1 h. Beads were washed four times with wash buffer (50 mM Tris pH 7.4, 300 mM NaCl, 0.5% NP-40, 0.5 mM PMSF, and protease inhibitor cocktail). Protein complexes were eluted by boiling in 50 µL 1× Laemmli Sample Buffer (Bio-Rad) at 100 °C for 10 min and chilled on ice for 5 min. DNase I treated IP was performed under the same conditions, with 25 U/mL DNase I added during antibody incubation. For western blotting, denatured samples were resolved on 4-15% Mini-PROTEAN TGX precast gels (Bio-Rad) and transferred to PVDF membranes using the Trans-Blot Turbo system (Bio-Rad). Membranes were blocked in 3% BSA in PBST (PBS containing 0.1% Tween-20) for 30 min at room temperature and then incubated overnight at 4°C with primary antibodies: anti-FLAG (CST, Cat. #14793; 1:2,000), anti-HA (CST, Cat. #2367T; 1:2,000), anti-BACH2 (Proteintech, Cat. #27635-1-AP; 1:2,000), and anti-MAFK/F/G (Santa Cruz, Cat. #sc-166548; 1:1,000). After three washes with PBST, membranes were incubated with HRP-conjugated anti-mouse or anti-rabbit secondary antibodies (1:5,000) and developed using ECL substrate (Bio-Rad). Signals were visualized using a ChemiDoc imaging system (Bio-Rad).

### Silver staining and in vitro native-PAGE assays

Recombinant human BACH2 (C-Myc/DDK-tagged, 92.4 kDa) and NRF2 (N-His-tagged, 67.6 kDa) were purchased from OriGene; MafK (N-His-tagged, 19.7 kDa) was purchased from ProSpec. Protein purity was assessed by SDS-PAGE followed by silver staining using the Pierce Silver Stain for Mass Spectrometry Kit (Thermo Fisher Scientific) before use in subsequent biochemical assays.

For *in vitro* analysis of the NRF2-BACH2 complex, recombinant BACH2, NRF2, and MafK proteins were mixed under the indicated conditions in binding buffer (10 mM HEPES, pH 7.5; 20 mM KCl; 1 mM MgCl_2_; 1 mM DTT) and incubated at room temperature for 25 min. Samples were then combined with 6× Native Sample Buffer (600 mM Tris-HCl, 50% glycerol, 0.02% bromophenol blue) and directly loaded onto 4-20% Mini-PROTEAN^®^ TGX™ precast gels (Bio-Rad) and run in Tris/Glycine electrophoresis buffer (Bio-Rad) at 4°C. Gels were subjected to standard western blotting procedures and were analyzed by immunoblotting with anti-BACH2 (Proteintech; Cat. #27635-1-AP) and anti NRF2 (CST; Cat. #12721T) antibodies at 1:2,000 dilution.

### Protein pull-down assay

Recombinant His-tagged NRF2 (5 pmol) or His-tagged MafK (5 pmol) was immobilized on 20 µL HisPur™ Ni-NTA Resin (Thermo Scientific) in binding buffer (50 mM Tris-HCl, pH 7.4, 150 mM NaCl, 20 mM imidazole, 0.05% NP-40, and 0.5 mM PMSF) for 1 h at 4 °C with rotation. After two washes with binding buffer, purified recombinant BACH2-MYC/DDK (10 pmol) was added and incubated for 2 h at 4 °C. A control reaction containing BACH2-MYC/DDK incubated with Ni-NTA Resin was performed in parallel to assess nonspecific binding of BACH2 to the Ni-NTA resin. Beads were washed three times with a wash buffer (50 mM Tris-HCl, pH 7.4, 300 mM NaCl, 20 mM imidazole, 0.05% NP-40, and 0.5 mM PMSF) of the same composition, and bound proteins were eluted by boiling in a 1× SDS sample buffer. Eluted proteins were analyzed by immunoblotting with anti-BACH2 (Proteintech; Cat # 27635-1-AP) and anti-His (Proteintech; Cat. # 10001-0-AP) antibodies at 1:2,000 dilution.

### Chromatin immunoprecipitation (ChIP)

ChIP was performed using chromatin prepared from 5×10^6^ differentiated erythroid progenitor cells derived from primary human CD34^+^ cells. On day 8 of erythroid differentiation, cells were cross-linked with 1% formaldehyde (Pierce Life Technologies, 28906) and quenched with glycine. Chromatin was processed using the truChIP Chromatin Shearing Kit (Covaris, 520127) according to the manufacturer’s protocol. Lysates were sonicated with an E220 sonicator (Covaris, 500239) to generate DNA fragments of 300–500 bp. Sheared chromatin was precleared with 15 µL Dynabeads Protein G (Invitrogen) supplemented with 100 µg BSA and 100 µg ssDNA. Precleared lysates were incubated overnight at 4 °C with 5 µg anti-NRF2 antibody (Abcam, ab62352) or normal rabbit IgG control (CST, 2729S). Beads were sequentially washed with 600 µL lysis buffer, 600 µL high-salt wash buffer (1% Triton X-100, 0.1% sodium deoxycholate, 50 mM Tris-HCl pH 8.0, 0.5 M NaCl, 5 mM EDTA), 600 µL LiCl immune complex wash buffer (0.25 M LiCl, 0.5% Igepal, 0.5% sodium deoxycholate, 10 mM Tris-HCl pH 8.0, 1 mM EDTA), followed by two washes with 600 µL TE buffer (10 mM Tris-HCl pH 8.0, 1 mM EDTA) at 4 °C. The complexes were eluted in 200 µL freshly prepared elution buffer (1% SDS, 0.1 M NaHCO_3_) with rotation at room temperature for 15 min. Reverse cross-linking was performed by adding 8 µL 5 M NaCl and incubating at 65 °C for 4 h, followed by the addition of 4 µL 0.5 M EDTA and 10 µL proteinase K (10 mg/mL) and incubation at 55 °C for 2 h. DNA was purified by phenol/chloroform extraction and ethanol precipitation with 20 µg yeast tRNA as a carrier. The DNA pellets were dissolved in 50 µL ddH_2_O for qRT-PCR analysis. Primer sequences are provided in **Supplementary Table 11**.

### Electrophoretic Mobility Shift Assay (EMSA)

Oligonucleotides used as IRDye-700 labeled probes are listed in **Supplementary Table 11**. The sense strand of each probe was synthesized with a 5′ IRDye 700 label (IDT) and annealed to the corresponding unlabeled antisense strand by heating at 100 °C for 5 min followed by slow cooling to room temperature. Unlabeled probes used for cold competition assays were prepared in the same manner. Nuclear extracts from K562 cells overexpressing either Flag-NRF2 or Flag-BACH2 were isolated using the NE-PER Nuclear and Cytoplasmic Extraction Kit (Thermo Scientific) according to the manufacturer’s instructions. For binding reactions with wild-type probes, varying amounts of nuclear extract (0.5-6 µg) were incubated with 5 nM IRDye-700 labeled DNA probes in binding buffer (10 mM Tris-HCl, pH 7.5; 50 mM KCl; 10 mM MgCl_2_; 3.5 mM DTT; 0.25% Tween-20; 50 ng/µL poly dI:dC) for 25 min at room temperature. For cold competition assays, 1 µM annealed unlabeled probe was added after the labeled probe and incubated for an additional 10 min. To compare binding affinity between NRF2 or BACH2 and edited probe variants, 4 µg of nuclear extract was incubated with 5 nM IRDye-700 labeled DNA probes in a binding buffer for 20 min at room temperature. Following incubation, samples were mixed with 10x Orange Loading Dye (LI-COR Biosciences) and resolved on Novex™ TBE Gels 4-20% (Invitrogen) in 0.5x TBE buffer at 150 V for 50 min. Gels were briefly rinsed in ddH_2_O and immediately imaged using an Odyssey CLx imager. Quantification of image was performed using the Image Studio software (LI-COR Biotech, v 6.1.0.79).

### Immunofluorescence and RNA In Situ hybridization

Immunofluorescence was performed as described previously with minor modifications. Briefly, 3 × 10^5^ cells were collected, washed once with DPBS, and resuspended in 0.1% BSA in DPBS. Cells were cytospun onto poly-D-lysine–coated German coverslips (Neuvitro Corporation) at 300 rpm for 4 min and fixed with 4% paraformaldehyde (Electron Microscopy Sciences, 15714) for 15 min at room temperature. Fixed cells were permeabilized with 0.5% Triton X-100 for 5 min on ice and blocked with 1% BSA in PBS for 30 min at room temperature. Cells were then incubated overnight at 4 °C with mouse anti-NRF2 (Santa Cruz, sc-365949, 1:50) and rabbit anti-BACH2 (CST, Cat.#80775, 1:50) antibodies. After primary incubation, AF647-conjugated goat anti-mouse IgG and AF594-conjugated goat anti-rabbit IgG secondary antibodies (1:1000) were applied for 1 h at room temperature in the dark. Nuclei were counterstained with DAPI for 1 min at room temperature. Coverslips were mounted on glass slides and sealed. Images were acquired using a Leica TCS SP8 confocal microscope.

For co-staining of the primary transcripts of *HBG1/2* (*pre-HBG1/2*) and NRF2, RNA FISH was performed first, followed by immunofluorescence. Briefly, cells were fixed with 4% paraformaldehyde for 15 min, permeabilized in 0.5% Triton X-100 containing 2 mM Ribonucleoside Vanadyl Complex (NEB), and dehydrated in 75% ethanol overnight. Cells were then incubated with 300 ng denatured Dig-labeled FISH probes in hybridization buffer (50% formamide in 2× SSC) at 50 °C overnight. After hybridization, cells were washed twice with Sal I buffer (50% formamide, 0.1× SSC, 0.1% SDS) and incubated with sheep anti-digoxigenin (Roche, 11333089001, 1: 400) for 1 h at room temperature. Following two washes with Sal II buffer (2× SSC containing 8% formamide), cells were incubated with mouse anti-NRF2 (Santa Cruz, sc-365949, 1:50) overnight at 4 °C. After two additional washes with Sal II buffer, cells were incubated with Cy3-conjugated donkey anti-sheep IgG (1:1000) and AF647-conjugated goat anti-mouse IgG secondary antibodies (1:1000) for 1 h at room temperature in the dark. Nuclei were counterstained with DAPI. Coverslips were mounted on glass slides and sealed. Images were acquired using a Leica TCS SP8 confocal microscope.

DIG-labeled *pre-HBG1/2* RNA probes were synthesized using the DIG RNA Labeling Mix (Roche, 11277073910) and T7 RNA Polymerase (Promega) according to the manufacturer’s instructions. DNA templates containing a T7 promoter were generated by PCR using *HBG1/2* intron 2 primers (**Supplementary Table 11**). In vitro transcription (IVT) was performed at 37 °C for 2 h using T7 RNA polymerase in the presence of the DIG RNA Labeling Mix, followed by DNase I treatment to remove the DNA template. RNA probes were purified using Monarch RNA Cleanup Columns (NEB) and analyzed by RNA gel electrophoresis to assess probe integrity and fragment size.

### Measurement of NRF2 and pre-*HBG1/2* transcription foci size

For quantifying NRF2 or *pre-HBG1/2* transcription foci in K562 cells, HSPCs, and differentiated erythroid cells, cells were subjected to IF or IF-FISH as described above. Images were acquired on a Leica TCS SP8 confocal microscope with z-stack acquisition. Raw images were imported into Fiji/ImageJ for analysis. Each channel was separated, and a maximum-intensity z-projection was generated. When necessary, background subtraction and linear contrast adjustment were applied uniformly across samples. NRF2 and *pre-HBG1/2* signals were then segmented using an optimal threshold. Regions of interest (ROIs) were defined based on the DAPI signal. Individual cells were analyzed using the “Analyze Particles” tool in Fiji/ImageJ (default settings: size 0-infinity, circularity 0.00-1.00). A total of 10-30 cells were quantified per condition, and the results were plotted in Prism 10.

### Colocalization analysis of NRF2 and pre-*HBG1/2*

Colocalization between NRF2 and *pre-HBG1/2* was quantified using Manders’ overlap coefficients (M1 and M2) implemented in the JaCoP plugin in Fiji/ImageJ. Images were acquired on a Leica TCS SP8 confocal microscope with z-stack acquisition. Raw images were imported into Fiji/ImageJ, channels were separated, and a maximum-intensity z-projection was generated for analysis. When necessary, background subtraction and linear contrast adjustment were applied uniformly across samples. Individual cells were segmented based on DAPI. Manders’ coefficients M1 and M2 were calculated following the optimal threshold to determine the fraction of signal from channel A overlapping with channel B, and vice versa. A total of 10-30 cells were quantified per condition, and the results were plotted in Prism 10.

### RNA-seq data processing

All RNA-seq data were aligned to the human reference genome hg38 using STAR^80^ with the parameters –runMode alignReads –chimSegmentMin 20. The resulting alignments were sorted, and unmapped reads were removed using SAMtools (v1.21) with option -F 516. Gene-level read counts were then generated by featureCounts^81^ using the human reference gene annotation. Differentially expressed genes (DEGs) were identified with DESeq2^82^ using the Wald test, applying thresholds of p < 0.05 and fold change > 1.4 (**Supplementary Table 10**)

### CUT&RUN and library preparation and sequencing

Cleavage Under Targets and Release Using Nuclease (CUT&RUN) experiments were carried out using CUTANA™ ChIC/CUT&RUN Kit, with slight modifications as described previously^83^. Briefly, ~500,000 cells were washed in a buffer containing digitonin (0.01% digitonin) and bound to 10 µl of activated Concanavalin A beads. Bead-bound cells were incubated overnight at 4°C with 0.5 µg NRF2 primary antibody (Sigma, SAB5700720), 0.5 µg NFE2 primary antibody (Sigma, HPA001914), or 0.5 µg BACH2 antibody cocktail. The BACH2 antibody cocktail consisted of three rabbit polyclonal anti-BACH2 antibodies targeting distinct epitopes (Sigma, HPA058384, aa 263-349; Sigma, HPA051384, aa 169-263; Abcam, ab226394, aa 750-841), premixed at equal concentrations. After washing, cells were incubated with pA/G-MNase for 1h at 4C and targeted chromatin digestion initiated by the addition of 100 mM CaCl_2_ and allowed to proceed for 30 min at 0°C, at which time stop buffer containing E. coli-Spike-in DNA was added. Incubate the reaction at 37°C for 30min, to release chromatin fragments and then purified by phenol-chloroform extraction followed by ethanol precipitation.

CUT&RUN DNA library preparation was carried out using The NEBNext Ultra II DNA Library Prep Kit as described previously^21^. Briefly, 15ng of CUT&RUN DNA were treated with endprep module at 20 °C for 30 min and 50 °C for 1 h to reduce the melting of short DNA. Ligation was performed by adding 1 pmol of NEB adapter and ligation mix and incubated at 20 °C for 15 min. To clean up the reaction, add 1.75× volume of SPRIselect beads (Beckman Coulter) to capture short ligation products. PCR amplification was performed for 12 cycles. The resulting libraries were purified with 1.2× volume of SPRIselect beads then analyzed and quantified by Qubit and Tapestation. Libraries with different indexes were pooled, and Illumina paired-end sequencing was performed using Nextseq platform with NextSeq-1000 P2 Kit (75 cycles) (2 × 50bp, 6-bp index).

### CUT&RUN data processing and peak calling

All CUT&RUN sequencing data in this study were processed using CUT&RUNTool (https://bitbucket.org/qzhudfci/cutruntools/src/default/)^84^, which performs sequential steps of read trimming, alignment, peak calling, and motif analysis. First, paired-end sequencing reads were trimmed with Trimmomatic (v0.36)^85^ to remove adapter sequences from the 3′ end using the parameters: ILLUMINACLIP:2:15:4:4: true LEADING:20 TRAILING:20 SLIDINGWINDOW:4:15. To further eliminate residual adapter sequences (up to 6 bp) not removed by Trimmomatic, an additional trimming step was performed using Kseq (https://bitbucket.org/qzhudfci/cutruntools/src/master/). The cleaned reads were then aligned to the human reference genome hg38 by Bowtie2 (v2.5)^86^ with parameters --dovetail and --phred33. The resulting BAM files were sorted, indexed, and duplicate reads were marked using Picard (v3.4.0) MarkDuplicates. Finally, unmapped, unmated, and duplicate reads were removed with SAMtools (v1.21)^87^, generating high-quality alignments for downstream analyses. MACS^72^ version 2.2 was used to call peaks from the BAM file with narrowPeak setting, with *q*-value cutoff 0.01.

### CUT&RUN normalizing to E.coli Spike-in DNA

To ensure comparability across CUT&RUN data, we normalized the data using *E. coli* spike-in DNA. In addition to aligning reads to the human reference genome hg38, sequencing reads were separately aligned to the *E. coli* K12 MG1655 reference, and non-uniquely mapped reads were removed. For each CUT&RUN dataset, the number of uniquely aligned *E. coli* reads was quantified and normalized to the total number of uniquely aligned human hg38 reads. A normalization factor was then calculated so that the *E. coli* spike-in signal was equalized across all CUT&RUN data. This single scalar normalization ratio was applied using the --scaleFactor option in the deepTools (v3.5.5)^88^ bamCoverage function to generate normalized bigWig files for visualization and comparative analyses.

### In silico base-perturbation analysis of transcription factor binding

To evaluate the contribution of individual nucleotides to the binding affinity of BACH2, NRF2, and NFE2, we generated a total of 5 variant sequences, each containing a single in silico base substitution relative to the reference sequence GTTTGCCTTGTCAAGGCTAT. These sequences were analyzed using FIMO^89^ for motif scanning with a significance threshold of p < 0.05. For each sequence, we extracted the log-likelihood ratio score reported by FIMO for potential motif matches at each position. Comparison of these scores allowed us to estimate the effect of single-base perturbations on motif binding affinity.

---

**Extended Data Fig. 1.**
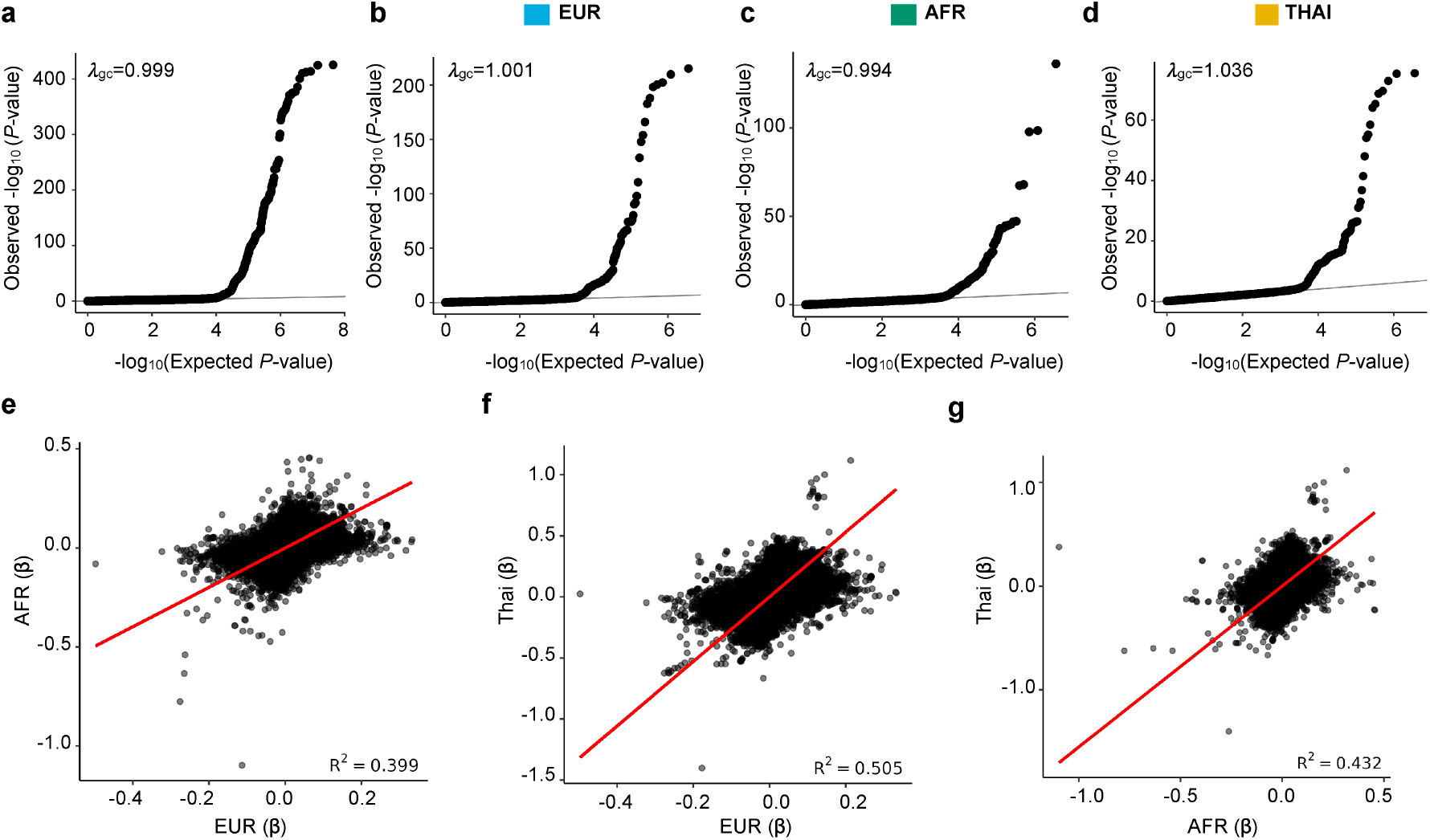
META GWAS and multi-ancestry meta-analysis QC and concordance across ancestry. **a-d**, Quantile-Quantile (QQ) plots for meta-analysis and ancestry-specific analyses using multi-ancestry meta-analysis (MAMA). LambdaGC values (λ_gc_) are annotated in each plot. **e-g**, Concordance of SNP effect sizes across ancestry groups in the MAMA. Correlations (R square value) are indicated on each plot.

**Extended Data Fig. 2.**
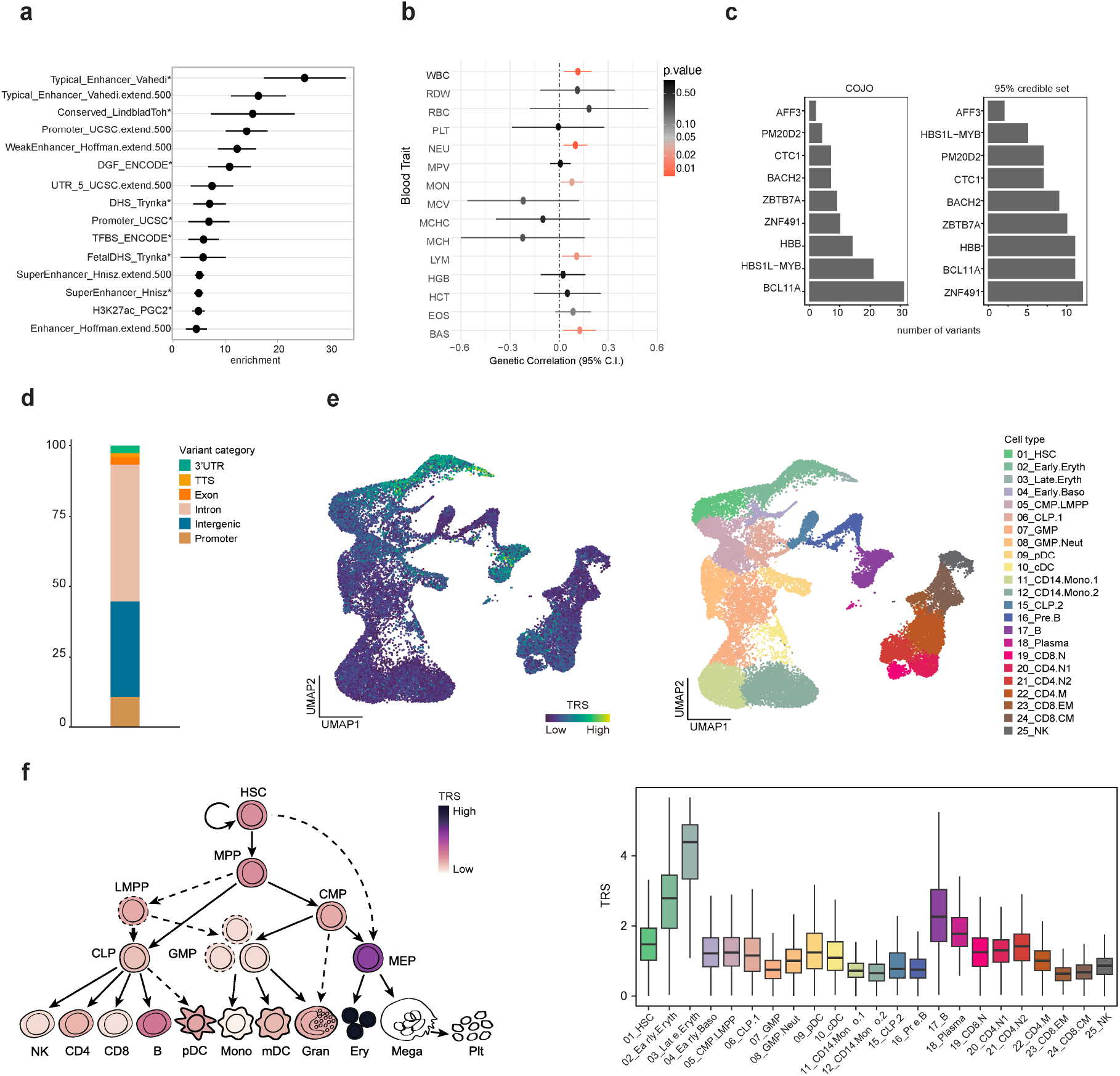
Heritability, genetic correlation, conditional and fine-mapping analysis. **a**, Heritability enrichments from LDAK for 64 functional categories in the BLD model; error bars represent the standard error. **b**, Genetic correlations between HbF (n=28,278) with various blood cell traits^25^ (n=746,667); error bars show the 95% confidence interval. **c**, Number of lead variants per locus in COJO analysis (left) and number of 95% credible set variants per locus in fine-mapping (right). **d**, Distribution of fine-mapped variants with PIP>0.001 across genomic annotations. **e**, UMAP projection of single cell ATAC-seq data colored by trait relevance score (TRS) from SCAVENGE (top) and cell types (bottom). **f**, Average TRS in major cell types along the hematopoietic lineage (top) and their distribution by UMAP clusters (bottom). Trait relevance scores (TRSs) for HbF levels across cell types along the hematopoietic hierarchy. Boxplots show the TRS for individual cells across cell types (from left to right: n = 1,469, n = 1,875, n = 142, n = 575, n = 2,715, n = 1,065, n = 2,505, n = 1,616, n = 823, n = 328, n = 2,286, n = 3,876, n = 822, n = 1,067, n = 1,822, n = 103, n = 688, n = 1,917, n = 1,667, n = 3,250, n = 542, n = 1,686, n = 949), with the median indicated by the center line, the 25th and 75th percentiles by the lower and upper bounds of the box, respectively, and whiskers extending to the minimum and maximum observations within 1.5 × the interquartile range (IQR) of the respective quartiles.

**Extended Data Fig. 3.**
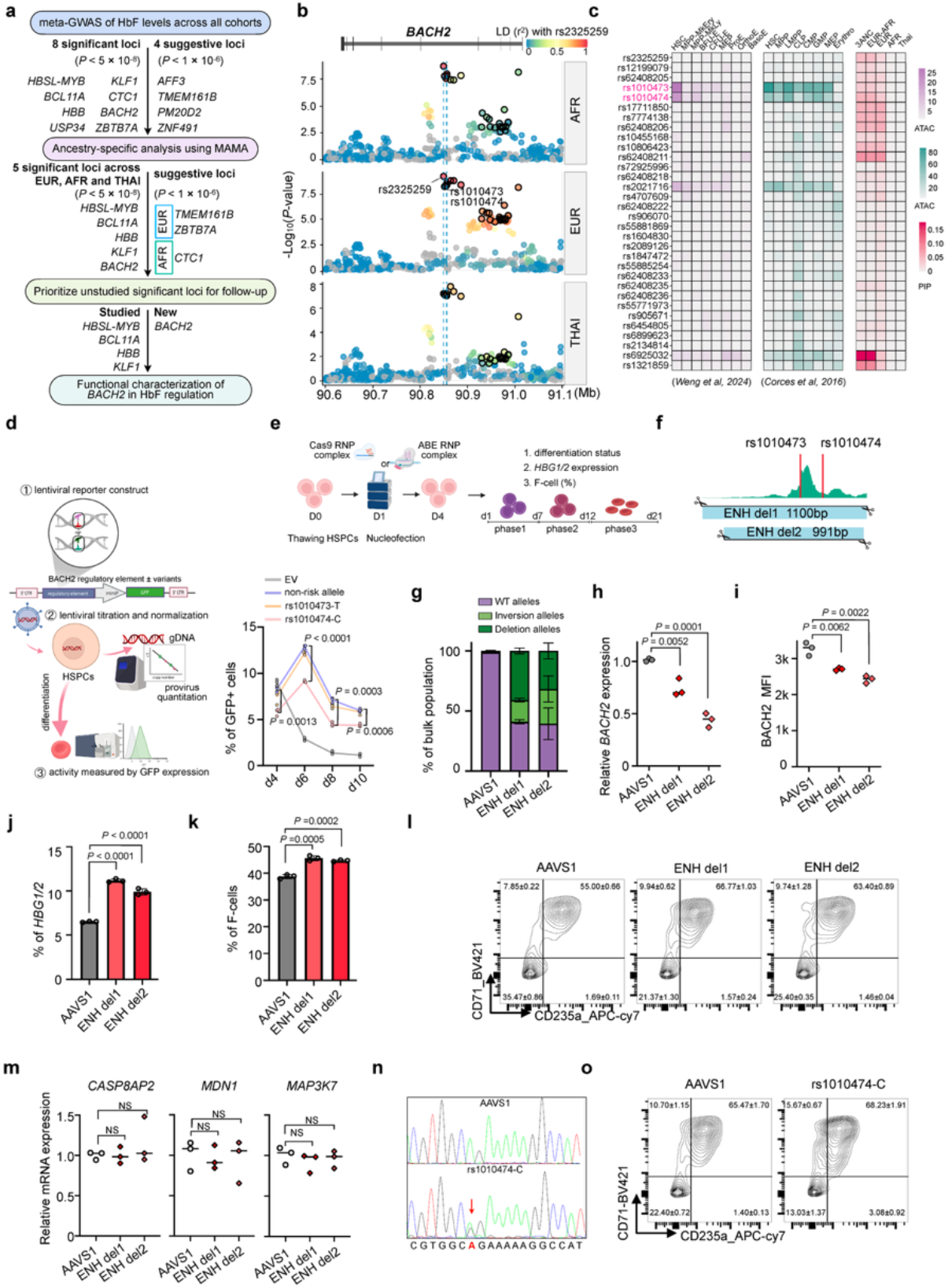
Variant rs1010474 and its harboring regulatory elements regulate *BACH2* and HbF expression. **a**, Study workflow for HbF GWAS and locus prioritization. Eight genome-wide significant loci were identified by meta-GWAS, five of which were replicated in ancestry-specific analyses. Among these, *BACH2* was the only previously unreported replicated locus and was prioritized for functional follow-up. **b**, Regional association plots showing MAMA signals (−log10*P*-value) for AFR, EUR, and Thai cohorts at the *BACH2* locus. The 32 candidate causal variants prioritized by Multi-SuSiE fine-mapping are indicated by circular dots with black borders. **c**, Functional prioritization of fine-mapped variants at the *BACH2* locus. Integrated heatmap of chromatin accessibility (ATAC-seq) and fine-mapping Posterior Inclusion Probabilities (PIP) for candidate causal variants. Normalized ATAC-seq signals across hematopoietic and erythroid lineages are shown for two independent datasets^90,91^ (purple and green scales). Statistical PIPs are displayed for the multi-ancestry meta-analysis (3ANC) and individual ancestry groups (red/blue scales). The variants rs1010473 and rs1010474 (highlighted in pink) are prioritized based on their high PIPs and precise overlap with prominent open chromatin regions across multiple progenitor subsets. **d**, Lentiviral reporter assay for *BACH2* enhancer variants during human erythroid differentiation. The *BACH2* regulatory element containing either non-risk or risk alleles was cloned into a reporter construct upstream of a minimal promoter driving GFP expression. Following lentivirus production and concentration, viral titers were normalized by quantifying integrated proviral copy numbers in K562 cells via qPCR. Titer-normalized viruses were then used to transduce primary human HSPCs, with regulatory activity assessed by GFP expression via flow cytometry during erythroid differentiation (left). Reporter activity was quantified as the percentage of GFP^+^ cells on day 4, 6, 8, 10 of erythroid differentiation (right). n = 3 independent experiments. **e**, Schematic of experimental design for *BACH2* regulatory element depletion by CRISPR/Cas9 and rs1010474-C base editing in human CD34^+^ HSPCs followed by erythroid differentiation and functional evaluation. **f**, Schematic of CRISPR/Cas9-mediated deletion of the *BACH2* regulatory element. Two pairs of guide RNAs (indicated by scissors) were designed to remove the open chromatin region (~0.6 kb, green peak). ENH del1 deleted 1,100 bp and ENH del2 deleted 991 bp within this region. The red line marks the positions of the variants. **g**, Enhancer perturbations were quantified by qPCR and shown as a percentage of total alleles in the bulk population. Plotted are wild-type alleles, inversions, and deletions in *AAVS1* control and deletion of the *BACH2* enhancer (ENH del1 and ENH del2). n = 3 independent experiments. **h-i**, Relative *BACH2* mRNA expression (**h**) and intracellular BACH2 protein levels (**i**) in human primary CD34^+^ HSPCs three days after deletion of *BACH2* enhancer, compared to *AAVS1* control. n = 3. **j**, Proportion of *HBG1/2* expression on day 13 of erythroid differentiation in *AAVS1* or *BACH2* ENH-deleted HSPCs. n = 3 independent experiments. **k**, Frequency of F-cells on day 13 of erythroid differentiation in *AAVS1* or *BACH2* ENH-deleted HSPCs. n = 3 independent experiments. **l**, Erythroid differentiation status in *AAVS1* control or *BACH2* ENH-deleted HSPCs at day 11 of erythroid differentiation, as assessed by surface expression of CD71 and CD235a. **m**, Relative expression of *MDN1, CASP8AP2*, and *MAP3K7* transcript in *AAVS1* control and *BACH2* Enhancer deleted HSPCs. n = 3 independent experiments. **n**, Editing efficiency at rs1010474-C following base editing on day 11 of erythroid differentiation, determined by Sanger sequencing. **o**, Erythroid differentiation status of *AAVS1* and rs1010474-C edited HSPCs at day 11 of erythroid differentiation. All data are presented as mean ± standard deviation (SD). Differences between groups were assessed using unpaired two-tailed t-tests. *P* > 0.05 was considered not significant (NS). Icons in 3 **d** and **e** are created in BioRender. Guo, C. (2026) https://BioRender.com/fde5g4u.

**Extended Data Fig. 4.**
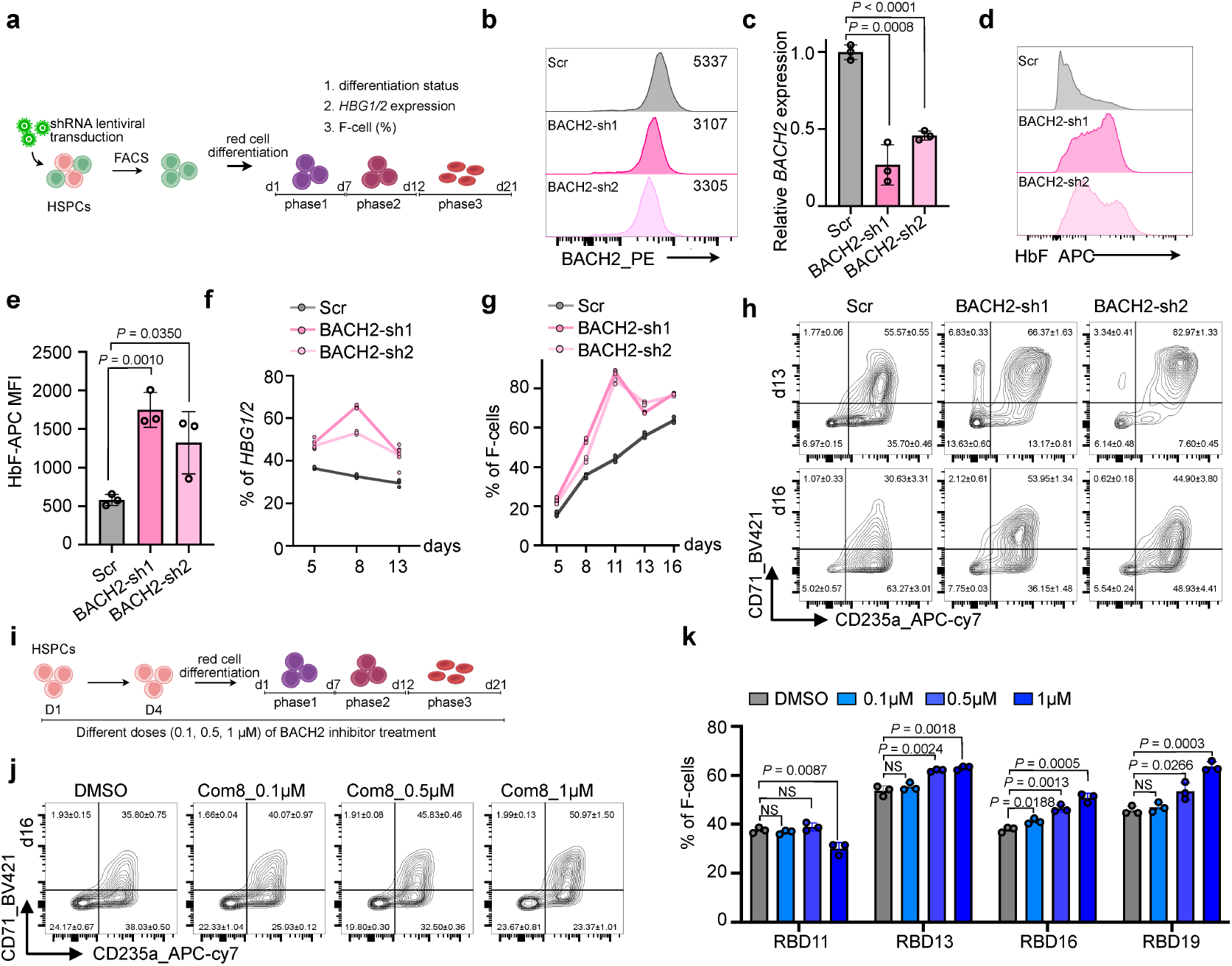
Knockdown or pharmacological inhibition of BACH2 promotes HbF expression. **a**, Schematic of experimental design for BACH2 knockdown in human CD34^+^ HSPCs followed by erythroid differentiation and functional evaluation. **b**, Representative histogram of BACH2 intracellular flow cytometric in bulk human primary CD34^+^ HSPCs four days after BACH2 knockdown. **c**, Relative *BACH2* mRNA expression on day 13 of erythroid differentiation in BACH2 shRNAs and Scr control. n = 3. **d-e**, Representative histogram (**d**) and quantification of MFI (**e**) of HbF intracellular flow on day 13 of erythroid differentiation in BACH2 shRNAs and Scr control. **f**, Proportion of *HBG1/2* expression on indicated time (d5, d8 and d13) of erythroid differentiation in BACH2 shRNAs and Scr control. n = 3. **g**, Frequency of F-cells on indicated time (d5, d8, d11, d13 and d16) of erythroid differentiation in BACH2 shRNAs and Scr control. n = 3. **h**, Erythroid differentiation status in BACH2 shRNAs and Scr control at indicated time (d13 and d16) of erythroid differentiation. **i**, Schematic of experimental design for BACH1/2 inhibitor (Comp8) treatment in HSPCs and during erythroid differentiation. **j**, Erythroid differentiation status following treatment with BACH1/2 inhibitor at day 16 of erythroid differentiation. **k**, Frequency of F-cells at indicated time points (d11, d13, d16, and d19) of erythroid differentiation following treatment with the BACH1/2 inhibitor (0.1-1μM). n = 3. All data are presented as mean ± standard deviation (SD). Differences between groups were assessed using unpaired two-tailed t-tests. *P* > 0.05 was considered not significant (NS).

**Extended Data Fig. 5.**
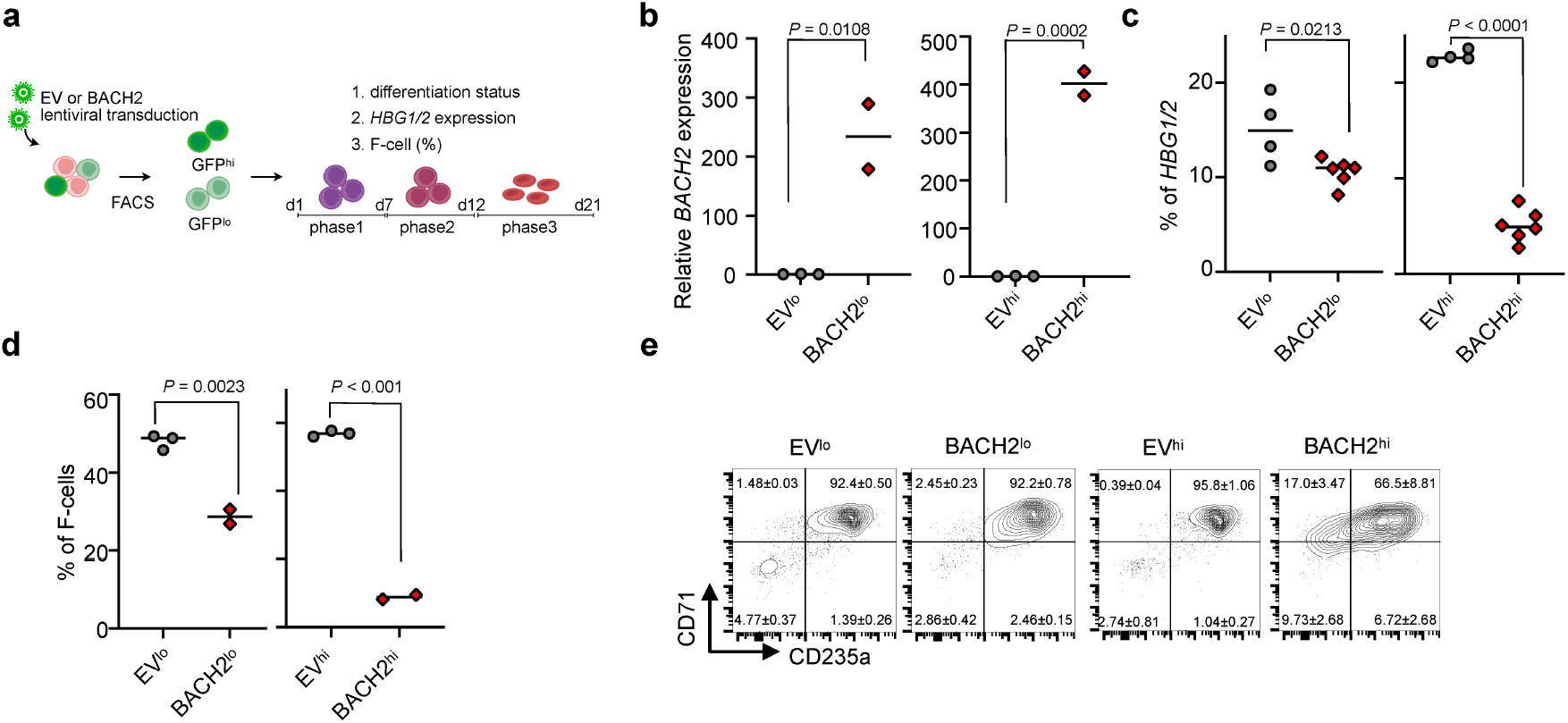
Overexpression of BACH2 inhibits HbF expression. **a**, Schematic of experimental design for BACH2 overexpression in human CD34^+^ HSPCs followed by erythroid differentiation and functional evaluation. **b**, Relative *BACH2* mRNA expression on day 11 of erythroid differentiation in BACH2^lo^ and BACH2^hi^ overexpressing HSPCs, compared with the corresponding EV control. n = 3 for control, n = 2 for overexpression, independent experiments. **c**, Proportion of *HBG1/2* expression on day 11 of erythroid differentiation in BACH2^lo^ and BACH2^hi^ overexpressing HSPCs. n = 4 for control, n = 6 for overexpression, independent experiments. **d**, Frequency of F-cells on day 11 of erythroid differentiation in BACH2^lo^ and BACH2^hi^ overexpressing HSPCs. n = 3 for control, n = 2 for overexpression, independent experiments. **e**, Erythroid differentiation status in BACH2^lo^ and BACH2^hi^ overexpressing HSPCs and corresponding EV controls at day 10 of erythroid cell differentiation. All data are presented as mean ± standard deviation (SD). Differences between groups were assessed using unpaired two-tailed t-tests.

**Extended Data Fig. 6.**
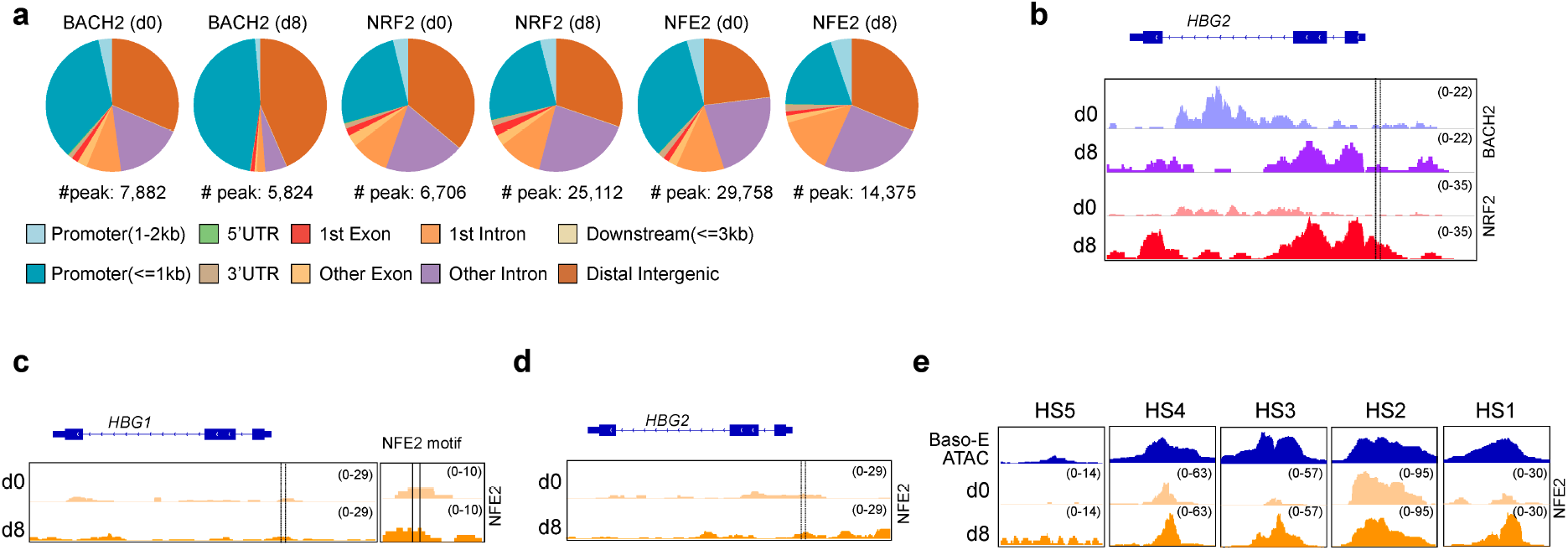
CUT&RUN profiling of BACH2 and NRF2 in human primary HSPCs and differentiated erythroid cells. **a**, Genome-wide distribution of BACH2 and NRF2, NFE2 CUT&RUN peaks in human CD34^+^ HSPCs (d0) and day 8 (d8) of erythroid differentiation. The number of peaks identified from CUT&RUN is indicated at the bottom. **b**, BACH2 or NRF2 occupancy at the *HBG2* promoter on day 0 and day 8 of erythroid differentiation. **c-d**, NFE2 occupancy at the *HBG1* or *HBG2* gene on d0 and d8 of erythroid differentiation. Insets highlight the NFE2 consensus motif at *HBG1* promoter. **e**, NFE2 occupancy at β-globin LCR on human day 0 and day 8 of erythroid differentiation.

**Extended Data Fig. 7.**
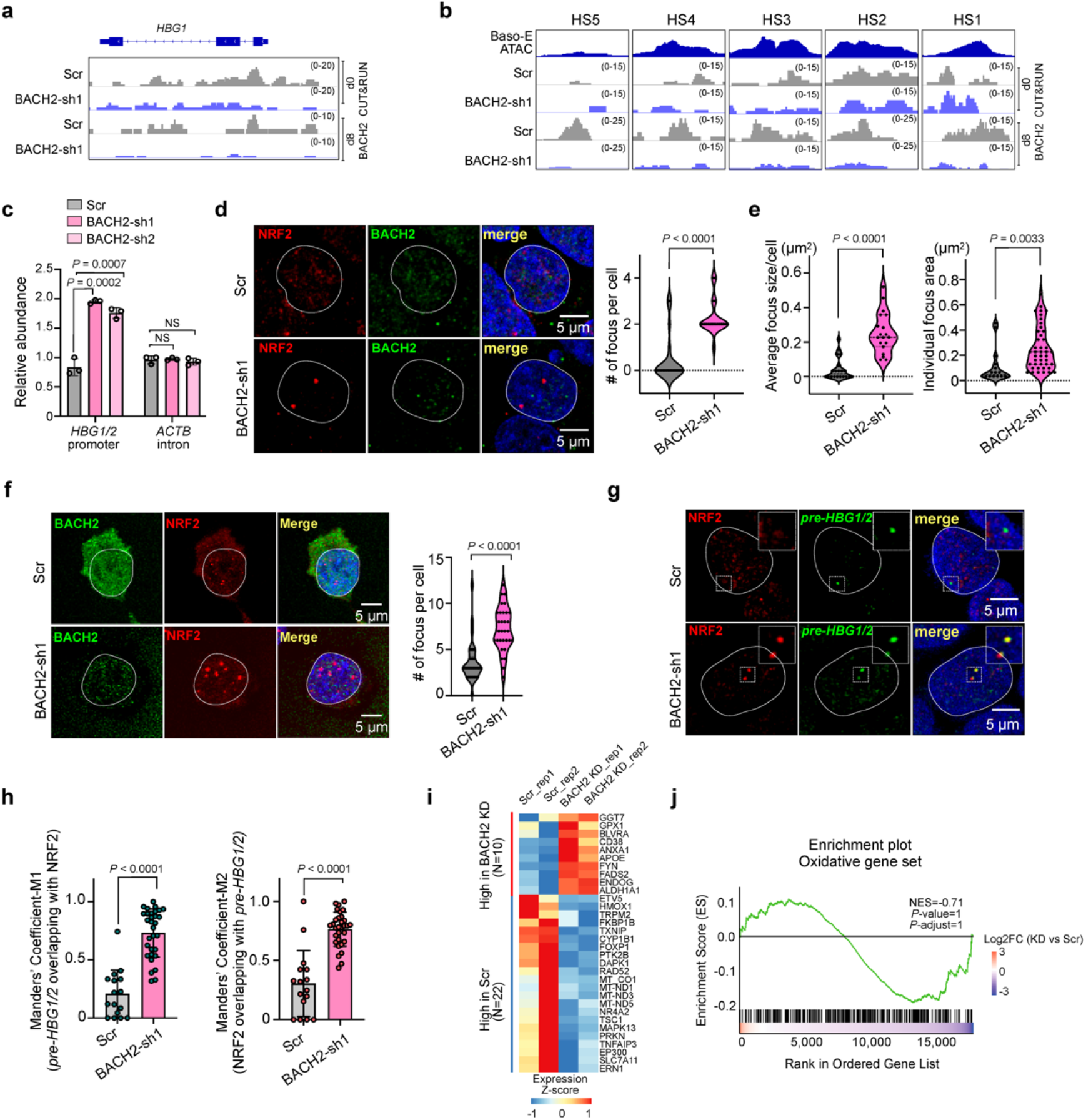
Knockdown of BACH2 enhances NRF2 accumulation at the -globin locus. **a**, BACH2 occupancy at the *HBG1* promoter on day 0 and day 8 of erythroid differentiation in Scr control and BACH2 knockdown HSPCs. **b**, BACH2 occupancy at LCR of β-globin on the day 0 and day 8 of erythroid differentiation in Scr control and BACH2 knockdown HSPCs. **c**, Relative NRF2 occupancy at *HBG1/2* promoter on day 8 of erythroid differentiation in BACH2 shRNAs and Scr control, revealed by ChIP-qPCR. ACTB intron serves as a negative control. n = 3 independent experiments. **d**, Co-staining of BACH2 and NRF2 in Scr control and BACH2 knockdown HSPCs (left). Quantification of NRF2 foci numbers in Scr control and BACH2 knockdown HSPCs (right). Each dot represents one cell; n = 20 cells for each condition. **e**, Quantification of average NRF2 focus size per cell (left) and individual NRF2 focus size (right) in Scr control and BACH2 knockdown HSPCs. n = 20 cells for each condition (left), n = 16 foci for Scr and n = 43 foci for BACH2 knockdown were measured (right). **f**, Co-staining of BACH2 and NRF2 in Scr control and BACH2 knockdown K562 cells (left). Quantification of NRF2 foci numbers in Scr control and BACH2 knockdown K562 cells (right). Each dot represents one cell; n = 30 cells for Scr, n = 33 cells for BACH2 knockdown. **g**, Co-staining of NRF2 and nascent *HBG1/2* transcripts (*pre-HBG1/2*) in Scr control and BACH2 knockdown HSPCs. **h**, Quantification of colocalization efficiency between NRF2 and *pre-HBG1/2* transcript by Manders’ coefficient on day 8 of differentiation in BACH2 shRNAs and Scr control. Each dot represents one cell. n = 16 cells for Scr, n = 31 cells for BACH2 knockdown. **i**, Differential expression of oxidative stress-related genes in Scr control and BACH2 knockdown erythroid cells. **j**, Gene enrichment analysis of oxidative stress-related genes in Scr control and BACH2 knockdown erythroid cells. All data are presented as mean ± standard deviation (SD). Differences between groups were assessed using unpaired two-tailed t-tests.

**Extended Data Fig. 8.**
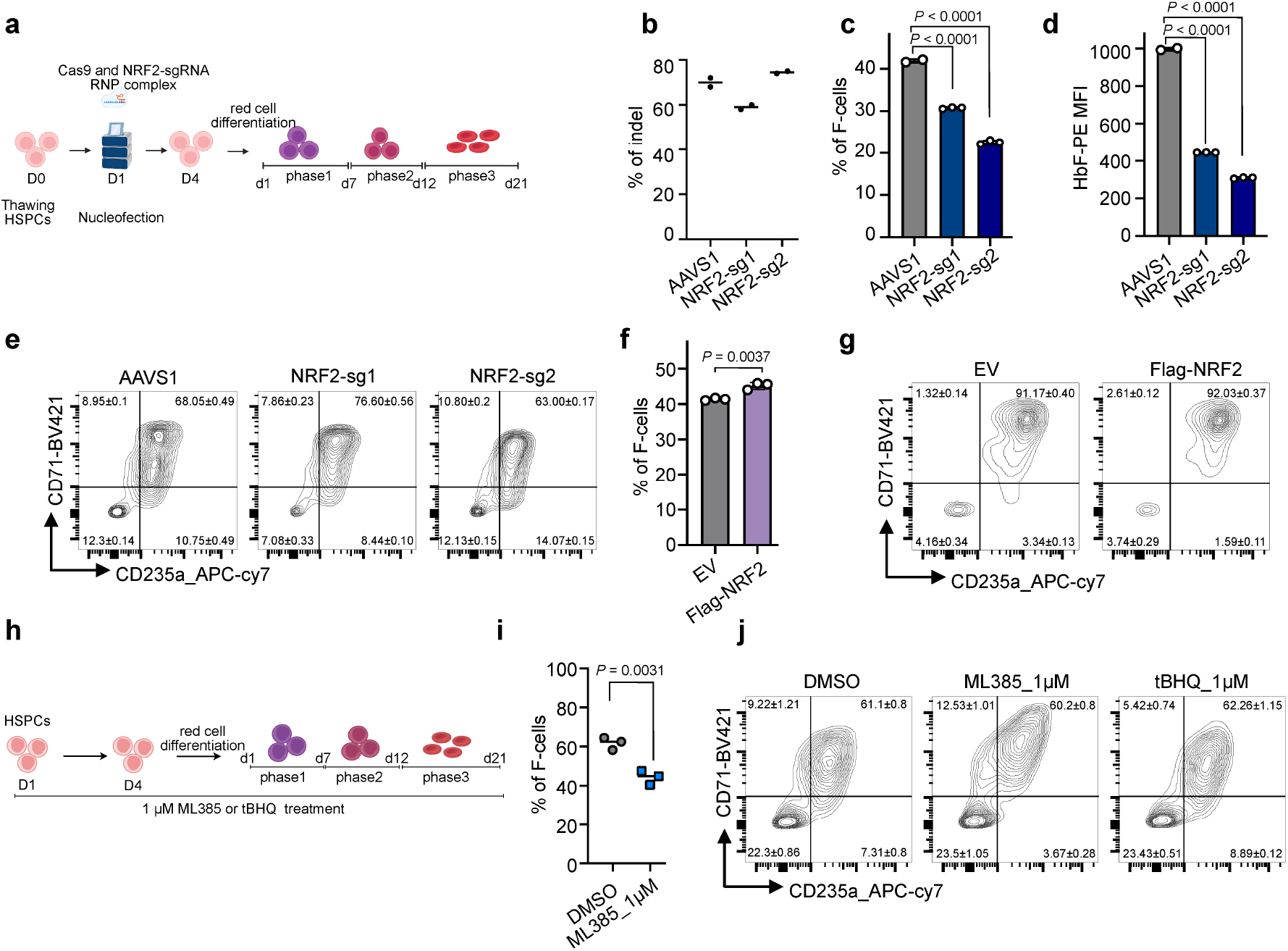
NRF2 functions as an activator to regulate HbF expression. **a**, Schematic of experimental design for CRISPR/Cas9-mediated NRF2 knockdown in human CD34^+^ HSPCs followed by erythroid differentiation and functional evaluation. **b**, ICE analysis of CRISPR/Cas9-mediated NRF2 knockdown efficiency on day 11 of erythroid differentiation. n = 2 independent experiments. **c**, Frequency of F-cells on day 11 of erythroid differentiation in *AAVS1* control and NRF2 knockdown HSPCs. n = 3 independent experiments. **d**, Quantification of MFI of HbF on day 11 of erythroid differentiation in *AAVS1* control and NRF2 knockdown HSPCs. n = 3 independent experiments. **e**, Erythroid differentiation status in *AAVS1* control and NRF2 knockdown HSPCs at day 16 of erythroid differentiation. **f**, Frequency of F-cells on day 11 of erythroid differentiation in EV control and Flag-tagged NRF2 overexpressing HSPCs. n = 3 independent experiments. **g**, Erythroid differentiation status in EV control and NRF2 overexpressing HSPCs at day 11 of erythroid differentiation. **h**, Schematic of experimental design for NRF2 inhibitor (ML385) and activator (tBHQ) treatment in human CD34^+^ HSPCs and during erythroid differentiation. **i**, Frequency of F-cells following ML385 treatment at day 11 of erythroid differentiation. n = 3 independent experiments. **j**, Erythroid differentiation status following treatment of ML385 or tBHQ at day 16 of erythroid differentiation. All data are presented as mean ± standard deviation (SD). Differences between groups were assessed using unpaired two-tailed t-tests. Icons in **a** are created in BioRender. Guo, C. (2026) https://BioRender.com/bvolp6h.

**Extended Data Fig. 9.**
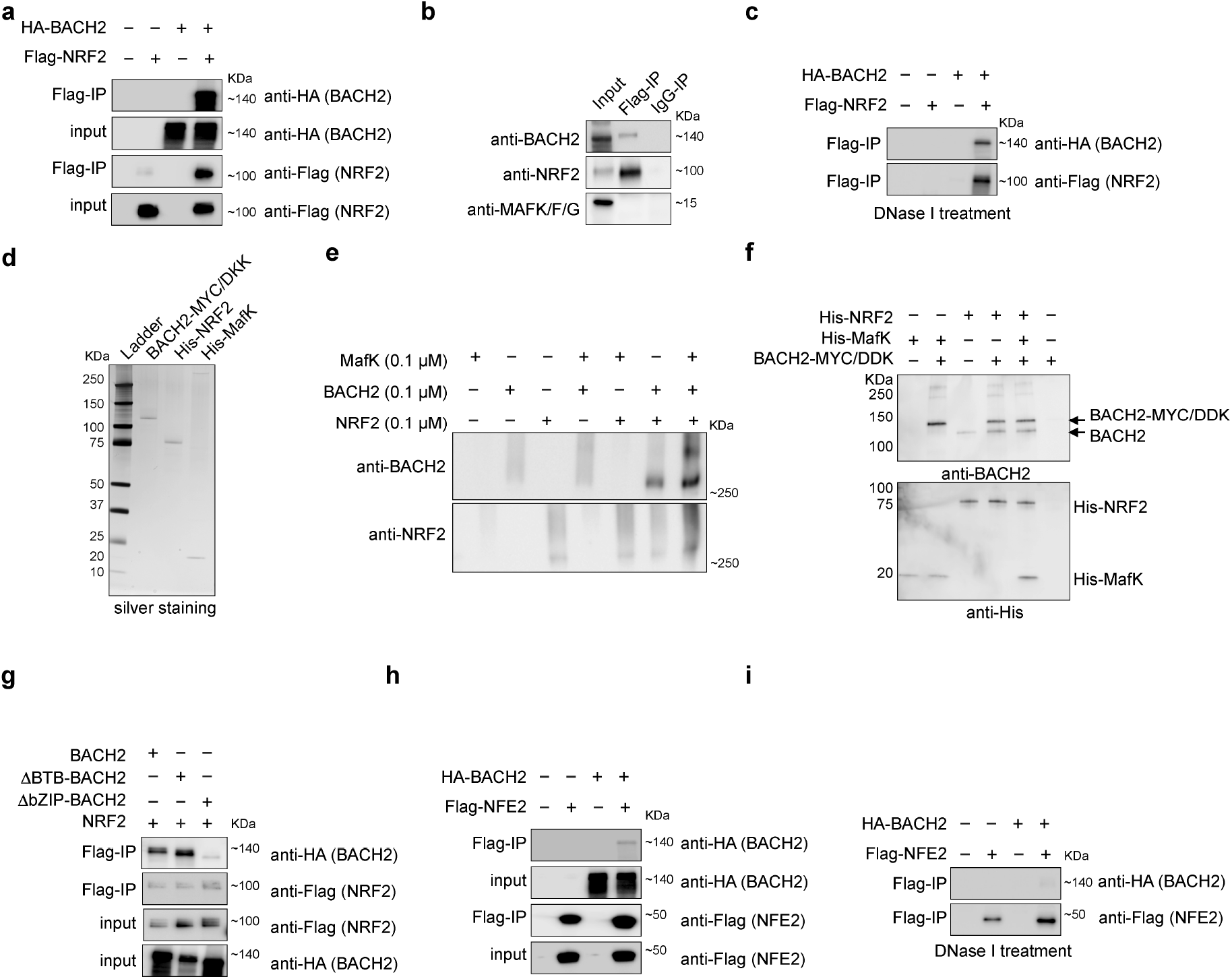
BACH2 specifically associates with NRF2, but not NFE2. **a**, NRF2 associates with BACH2. Co-immunoprecipitation (Co-IP) was performed using an anti-Flag antibody in HEK293 cells overexpressing Flag-tagged NRF2 and HA-tagged BACH2, followed by immunoblotting with an anti-HA antibody. **b**, NRF2 associates with endogenous BACH2 under stringent washing conditions. Co-IP was performed using anti-Flag antibody in K562 cells overexpressing Flag-tagged NRF2, followed by immunoblotting with antibodies against endogenous BACH2 and MAFK/F/G. **c**, NRF2 associates with BACH2 upon DNase I treatment. Co-IP was performed using anti-Flag antibody in HEK293 cells overexpressing Flag-tagged NRF2 and HA-tagged BACH2 in the presence of DNase I, followed by immunoblotting with an anti-HA antibody. **d**, Silver staining of recombinant proteins separated by SDS-PAGE. **e**, Native PAGE analysis of recombinant BACH2 and NRF2 proteins. Immunoblotting of the native gel showed co-migration of BACH2 and NRF2 at the same position, with or without MAFK. **f**, His-tagged protein pull-down assay using recombinant proteins. Purified proteins were subjected to His tag-mediated pull-down and analyzed by immunoblotting with anti-BACH2 (top) and anti-His (bottom) antibodies. **g**, NRF2 interacts with BACH2 through the bZIP domain. Co-IP was performed using an anti-Flag antibody in HEK293T cells overexpressing Flag-tagged NRF2 and full length or truncated HA-tagged BACH2, followed by immunoblotting with an anti-HA antibody. **h-i**, NFE2 does not associate with BACH2. Co-IP was performed using an anti-Flag antibody in HEK293 cells overexpressing Flag-tagged NFE2 and HA-tagged BACH2 (**h**) or after DNase I treatment (**i**), followed by immunoblotting with an anti-HA antibody.

**Extended Data Fig. 10.**
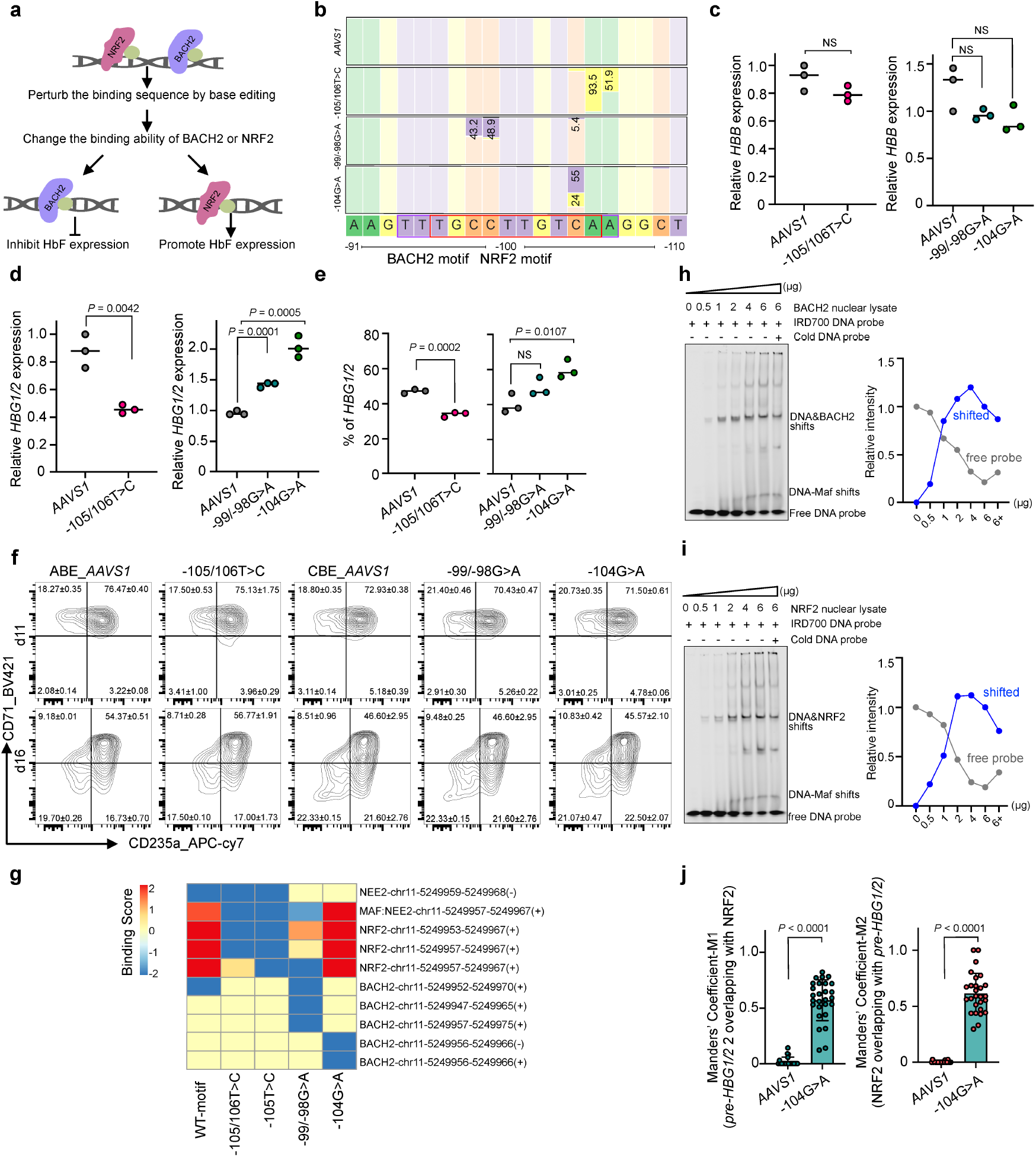
Editing of BACH2/NRF2 binding motifs in the -globin. **a**, Proposed model of base editing at BACH2 and NRF2 binding motifs in the -globin promoter. Editing of the *cis* elements alters the binding preference of BACH2 or NRF2, leading to differential regulation of HbF expression. **b**, Editing efficiency of BACH2 and NRF2 binding motifs in the -globin on day 11 of erythroid differentiation, revealed by CRISPResso analysis. **c**, Relative *HBB* expression on day 11 of erythroid differentiation in BACH2 and NRF2 binding motifs edited HSPCs, compared to *AAVS1* control. n = 3 independent experiments. **d**, Relative *HBG1/2* expression on day 11 of erythroid differentiation in BACH2 and NRF2 binding motifs edited HSPCs, compared to *AAVS1* control. n = 3. **e**, Proportion of *HBG1/2* expression on day 7 of erythroid differentiation in BACH2 or NRF2 motif edited HSPCs, compared to *AAVS1* control. n = 3 independent experiments. **f**, Erythroid differentiation status of *AAVS1* control and BACH2 and NRF2 motif edited HSPCs on day 11 and 16 of erythroid differentiation. **g**, BACH2 and NRF2 binding affinity by *in-silico* base-perturbation analysis. **h**, EMSA (left) and quantification (right) of BACH2 binding to the WT DNA probe. A titration of BACH2 nuclear lysate (0-6 μg) was used to show dose-dependent binding. Competition with excess unlabeled cold DNA probes in the final lane (marked as 6+ on the graph) demonstrates binding specificity. Indicated bands represent DNA-BACH2 super-shifts, shifts, and endogenous DNA-Maf complexes. **i**, EMSA and quantification as in (**h**), showing the dose-dependent binding of NRF2 nuclear lysate to the same DNA probe. Specific DNA-NRF2 shifted bands and endogenous DNA-Maf complexes are indicated. **j**, Quantification of colocalization efficiency between NRF2 and *pre-HBG1/2* transcript by Manders’ coefficient in *AAVS1* control and G^104^>A motif edited cells on day 8 of erythroid differentiation. Each dot represents one cell; n = 22 for *AAVS1*, n= 27 for G^104^>A edited. All data are presented as mean ± standard deviation (SD). Differences between groups were assessed using unpaired two-tailed t-tests.

## Notes

### Author Declarations

Ethics committee/IRB of Boston Children's Hospital gave ethical approval for this work.

### Summary of Updates

Revised and updated analyses provided, along with corrections to the genetic associations.

