## Supplementary Notes for "Human genetics implicates a BACH2-NRF2 axis in fetal heamoglobin activation"

#### Cohort-specific data for the genome-wide association studies performed

#### European cohorts

The samples from Sweden (N=3,031), SardiNIA (N=6,305), and INTERVAL (N=11,004) are phenotyped by hemoglobin HPLC. GWAS were conducted independently within each cohort. Note that the sample size N here (and the “Sample Size” column in **Supplementary Table 1**) denotes the number of samples that passed QC filters and are subsets of all genotyped samples in the respective cohorts.

The SardiNIA cohort^1,2^ includes 6,305 unrelated individuals recruited from clinics in four towns in the Lanusei Valley of Sardinia, Italy. Ethical approval for the study was obtained from the IRBs of the Istituto di Neurogenetica e Neurofarmacologia (INN; Cagliari, Italy), MedStar Research Institute (intramural at the National Institutes of Aging, NIH), and the University of Michigan. Genotyping was performed using Affymetrix 10K and 500K arrays and imputed to the WGS data from the same population.

The Swedish cohort^3^ recruited participants from the Skåne county in southern Sweden. The study received ethical approval from the Lund University Ethical Review Board (Approval 2013/54). Hemoglobin HPLC was conducted on frozen blood samples for all donors. Genotyping was conducted using Illumina Omni-1 Quad BeadChips and subsequently imputed to the TOPMed reference panel. Variants were filtered by INFO >= 0.6, MAF >= 0.001, and MAC >= max(20, min(40, $\sqrt{N}$)) before being harmonized for the meta-analysis.

The INTERVAL cohort included around 12,000 generally healthy adult whole-blood donors recruited across 25 static centers of NHS Blood and Transplant (NHSBT) throughout England, UK. Ethical oversight was provided by the UK’s National Research Ethics Service (Approval #: 11/EE/0538) and the Cambridge (East) Research Ethics Committee. Samples were genotyped using the ThermoFisher UK Biobank array^4^ and imputed to a combined 1000 Genomes Phase 3 (1KG-Ph3) + UK10K reference panel. HbF levels were stepwise regressed onto covariates including age, age^2^, blood group, INTERVAL trial arm, recruitment center, recruitment date, assay run number, and the top 10 genetic principal components (PCs), which together explained 32.6% of the HbF level variation. Residuals were then rank inverse normal transformed for GWAS. After removing duplicate samples and samples of non-European ancestry, missing covariates, or missing HbF levels, 11,004 samples remained.

Variants within the INTERVAL cohort were initially in GRC37 and filtered by INFO>=0.4 and MAC>=40. GWAS was then performed by BOLT-LMM v2.3 on autosomal variants under an additive model. Bi-allelic single-nucleotide variants with INFO >= 0.6 and MAF >= 0.001 were further selected for downstream meta-analysis.

The samples from GTEx^4,5^ (v8; N=670) and BIOS^6^ (Biobank-based Integrative Omics Study, part of eQTLgen) consortia have transcriptomic data, and therefore have RNA expression levels of *HBG1*, *HBG2*, and *HBB* gene expression available as proxies for HbF phenotypes. More specifically, the ratio of (*HBG1* + *HBG2*)/*HBB* was inverse-normal transformed as the phenotype used for GWAS.

The samples from the GTEx cohort were from diseased donors and shipped to the GTEx Laboratory Data Analysis and Coordination Center (LDACC) at the Broad Institute. Detailed QC information regarding WGS and expression data can be found in GTEx Consortium (V8)^5^. There were 670 participants with genotypes from WGS and whole blood samples that passed QC. These samples were selected from the TPM expression matrix (GTEx_Analysis_2017-06-05_v8_RNASeQCv1.1.9_gene_tpm[.gct.gz](http://.gct.gz), available for download at <https://gtexportal.org>), from which the ratio (*HBG1* + *HBG2*) /*HBB* was computed and inverse-normal transformed. The GWAS was performed with TensorQTL^7^ following variant-filtering criteria for trans-QTL mapping in GTEx Consortium (V8)^5^, using 5 in-sample genotype PCs, 60 PEER factors, sequencing platform (HiSeq 2000 or HiSeq X), LC protocol (PCR-based or PCR-free), and sex as covariates. Variants with in-sample MAF >= 0.05 were retained in the analysis.

Within the BIOS Consortium, we assessed samples from the Leiden Longevity Study^8,9^ (LLS, N=392), LifeLines DEEP^10^ (LL, N=738), and Rotterdam Study^10,11^ (RS; N=742). GWAS was conducted separately for each of them.

The LLS cohort consists of long-living Dutch participants whose biological samples were drawn during home visits. The study was approved by the Ethical Committee of the Leiden University Medical Center. The samples in the LL cohort were collected at the LifeLines Research Site in Groningen, The Netherlands. Ethical approval was granted by the Ethics Committee of the University Medical Centre Groningen. The RS cohort collected samples in the Ommoord district in the city of Rotterdam, The Netherlands. Ethical oversight was provided by the Medical Ethics Committee of the Erasmus Medical Center and the IRB of the Netherlands Ministry of Health, Welfare and Sports.

Genotyping for LLS, LL, and RS samples was done using Illumina Human660W-Quad and OmniExpress BeadChips, the HumanCytoSNP-12 BeadChip and the ImmunoChip (a customized Illumina Infinium array), and the Illumina 610K quad beadchip array, respectively. These chip-array data were subsequently imputed to the Haplotype Reference Consortium (HRC) using the Michigan imputation server. GWAS was then performed on each of these three cohorts separately on the University of Michigan Genome Center server using SAIGE v0.44.6.4, under the settings --minMAF 1e-4 --minInfo 0.6 --IsSparse TRUE --SPAcutoff 2 --LOCO FALSE --IsDropMissingDosage FALSE. Bi-allelic single-nucleotide variants with INFO >= 0.6, MAF >= 0.001, and MAC >= max(20, min(40, $\sqrt{N}$)) were retained for analysis.

#### Sickle cell disease (SCD) cohorts of African ancestry

We had three sets of GWAS performed on the Tanzania^12^ panel, the St. Jude Sickle Cell Clinical Research & Intervention Program (SCCRIP)/Baylor^13,14^ panel, and the TOPMed panel (including Walk-PHaSST^15^, OMG-SCD^16^, and REDS-III Brazil^17^ studies), respectively. All three panels consist of SCD individuals of African or genetically-inferred African ancestry.

The Tanzania cohort^12^, was based at Muhimbili National Hospital, Dar es Salaam, Tanzania, and approved by the Muhimbili University Research & Publications Committee (MU/RP/AEC/VOL.XIII). Samples were genotyped using the Illumina Human Omnichip 2.5 and imputed to the TOPMed reference panel. GWAS was performed by REGENIE v2.2.4, in which leave-one-chromosome-out (LOCO) predictions were generated during Step 1 on a subset of high-quality variants, before association testing was performed on the imputation dosage accordingly. We included age, sex, SCD treatment, and the top 10 PCs as covariates of the linear model, with sex and treatment as categorical variables. Among 1,843 samples with genotype data, 1,187 have non-missing phenotype and covariate data.

The Sickle Cell Clinical Research and Intervention Program (SCCRIP)^10,11,14^ cohort (N=584) was conducted at St. Jude Children’s Research Hospital, Methodist University Hospital, Our Lady of the Lake Children’s Hospital, OSF Healthcare Children’s Hospital of Illinois, and Novant Health Hemby Children’s Hospital. The SCCRIP Steering Committee and the IRBs of Baylor College of Medicine and St. Jude provided ethical oversight. Whole-genome sequencing was performed using Illumina HiSeq X10. Mapping (to GRCh38), variant calling, and QC details have been described previously^14^. In particular, PCs were generated by PC-AiR using bi-allelic variants in a combined panel of the samples of 327 study participants with CEU, YRI, CHB, and JPT samples from the 1000 Genomes Project. Long-range LD regions were excluded before PC calculation. After removing samples with missing data, 567 samples remain. GWAS was performed on variants with MAF > 0.01 using EMMAX^18^, a mixed-model association testing framework within the EPACTS suite that accounts for population structure and hidden relatedness. A genomic kinship matrix was generated from all genotyped cohort samples for relatedness adjustment, with which an unconditional analysis was run on square-root-transformed %HbF as the phenotype, using sex, cubic-spline-defined age, recruitment site location, and the top 10 PCs as covariates.

The Walk_PHaSST (TOPMed) cohort comprises 408 participants (dbGaP #: phs001514.v3.p1; Clinical Trial #: NCT00492531). The collaborating institutions were Children's Hospital of Pittsburgh of UPMC, Columbia University, Albert Einstein College of Medicine, National Heart & Lung Institute, UCSF Benioff Children's Hospital Oakland, Imperial College of London, Hammersmith Hospital, London, Howard University, University of Colorado, Denver, University of Illinois Chicago, Johns Hopkins University, and the National Heart, Lung, and Blood Insitute (NHLBI). The study was approved by the IRBs of UPitt, ColumbiaU, National Heart & Lung Institute, Imperial College London, HowardU, UColorado Denver, UIC, JHU, and NHLBI/NIH. Sequencing was conducted at the Baylor Genomics Center (HHSN268201500015C).

The OMG_SCD (TOPMed) cohort has 253 participants (dbGaP #: phs001608). The study sites were Duke University Medical Center and the University of North Carolina Chapel Hill Comprehensive Sickle Cell Centers. Ethical review was performed by the IRBs of Duke University and the University of North Carolina. Sequencing was carried out at the Baylor College of Medicine Human Genome Sequencing Center (HHSN268201600033I) using the Illumina HiSeq X10 platform.

The REDS-III Brazil (TOPMed) cohort includes 1,589 participants (dbGaP #: phs001468.v3.p1). The large-scale domestic and international collaboration involves blood centers, hospitals, a data coordinating center, and a central laboratory in the US, along with distinct programs in Brazil, China, and South Africa. The study was overseen by the REDS-III Observational Study Monitoring Board and the IRB of NHLBI. Sequencing was performed at the Baylor Genomics Center (HHSN268201600033I & HHSN268201500015C) using Illumina HiSeq X10.

Post imputation and QC samples from all TOPMed cohorts (dbGaP freeze 8 release) were pooled to perform a single GWAS on the TOPMed server. Sample PCs were obtained from dbGaP freeze 8 release samples and were produced by PC-Air. GWAS was performed in SAIGE using sex, age, and the top 10 PCs as covariates. Bi-allelic single-nucleotide variants with INFO >= 0.6, MAF >= 0.001, and MAC >= max(20, min(40, $\sqrt{N}$)) were retained for analysis.

#### HbF-screened Thai cohort

We recruited Thai participants at Siriraj Hospital at Mahidol University, Bangkok, Thailand, with ethics approval from the IRB of Siriraj Hospital at Mahidol University. Around 86,000 people from the general population had their HbF levels screened at the center, from which we selected individuals with HbF levels above 2% of total hemoglobin. The thalassemia status of all screened participants was examined at the hospital thalassemia study center, and those with severe forms of thalassemia were excluded from this study.

We randomly selected individuals from participants with normal HbF levels to make up a cohort of 1,474 individuals to genotype with Illumina Multi-Ethnic Global Array (MEGA). Among them, 147 high-HbF and 51 normal samples were further selected to have their whole genomes sequenced with Illumina HiSeq 4000 at the Broad Institute. Whole-genome genotypes of the remaining 1273 individuals were subsequently imputed based on the 198 WGS samples using IMPUTE2. Next, sample- and variant-level QCs were performed in Hail. Variants with DP >=10, GQ>=20, freq>=0.0001, and call_rate>0.9 were kept, while variants flagged for excess heterozygosity (“ExcessHet”) or low-confidence VQSR tranches (“VQSRTrancheSNP99.00to99.90+”, “VQSRTrancheINDEL99.90to99.95”, and “VQSRTrancheINDEL99.95to100.00” flags) were removed. Samples with call rates under 0.95 were removed, leaving 1,392 samples ready for GWAS. GWAS was performed with REGENIE using sex, age, and the top 10 PCs as covariates.

### Supplementary Acknowledgements

Molecular data for the TOPMed program was supported by the National Heart, Lung and Blood Institute (NHLBI). WGS for “NHLBI TOPMed: walk_PHaSST” (phs001514) was performed at the Baylor Genomics Center (HHSN268201500015C). WGS for “NHLBI TOPMed: Whole Genome Sequencing and Related Phenotypes in the Outcome Modifying Genes in Sickle Cell Disease Study” (OMG-SCD) (phs001608) was performed at the Baylor College of Medicine Human Genome Sequencing Center (HHSN268201600033I). WGS for “NHLBI TOPMed: REDS-III_Brazil” (phs001468) was performed at the Baylor Genomics Center (HHSN268201600033I & HHSN268201500015C). Core support including centralized genomic read mapping and genotype calling, along with variant quality metrics and filtering, was provided by the TOPMed Informatics Research Center (3R01HL-117626-02S1; contract HHSN268201800002I). Core support including phenotype harmonization, data management, sample-identity QC, and general program coordination, was provided by the TOPMed Data Coordinating Center (R01HL-120393; U01HL-120393; contract HHSN268201800001I).

We gratefully acknowledge the studies and participants who provided biological samples and data for TOPMed. The OMG-SCD study was administrated by Marilyn J. Telen, M.D. and Allison E. Ashley-Koch, Ph.D. from Duke University Medical Center, and the collection of the data set was supported by grants HL068959 and HL079915 from the National Heart, Lung, and Blood Institute (NHLBI) of the National Institute of Health (NIH).

We thank Dr. Mark Gladwin and the investigators of the Walk-PHasst study and the patients who participated in the study. We also thank the walk-PHaSST clinical site team: Albert Einstein College of Medicine: Jane Little and Verlene Davis; Columbia University: Robyn Barst, Erika Rosenzweig, Margaret Lee and Daniela Brady; UCSF Benioff Children's Hospital Oakland: Claudia Morris, Ward Hagar, Lisa Lavrisha, Howard Rosenfeld, and Elliott Vichinsky; Children's Hospital of Pittsburgh of UPMC: Regina McCollum; Hammersmith Hospital, London: Sally Davies, Gaia Mahalingam, Sharon Meehan, Ofelia Lebanto, and Ines Cabrita; Howard University: Victor Gordeuk, Oswaldo Castro, Onyinye Onyekwere, Vandana Sachdev, Alvin Thomas, Gladys Onojobi, Sharmin Diaz, Margaret Fadojutimi-Akinsiku, and Randa Aladdin; Johns Hopkins University: Reda Girgis, Sophie Lanzkron and Durrant Barasa; NHLBI: Mark Gladwin, Greg Kato, James Taylor, Wynona Coles, Catherine Seamon, Mary Hall, Amy Chi, Cynthia Brenneman, Wen Li, and Erin Smith; University of Colorado: Kathryn Hassell, David Badesch, Deb McCollister and Julie McAfee; University of Illinois at Chicago: Dean Schraufnagel, Robert Molokie, George Kondos, Patricia Cole-Saffold, and Lani Krauz; National Heart & Lung Institute, Imperial College London: Simon Gibbs. Thanks also to the data coordination center team from Rho, Inc.: Nancy Yovetich, Rob Woolson, Jamie Spencer, Christopher Woods, Karen Kesler, Vickie Coble, and Ronald W. Helms. We also thank Dr. Yingze Zhang for directing the Walk-PHasst repository and Dr. Mehdi Nouraie for maintaining the Walk-PHasst database and Dr. Jonathan Goldsmith as a NIH program director for this study. Special thanks to the volunteers who participated in the Walk-PHaSST study. This project was funded with federal funds from the NHLBI, NIH, Department of Health and Human Services, under contract HHSN268200617182C. This study is registered at www.clinicaltrials.gov as NCT00492531. Detailed description of the study was published in Blood, 2011 118:855-864, Machado et al. "Hospitalization for pain in patients with sickle cell disease treated with sildenafil for elevated TRV and low exercise capacity".

The fetal hemoglobin measurements in the INTERVAL consortium were funded by Biogen. Participants in the INTERVAL randomised controlled trial were recruited with the active collaboration of NHS Blood and Transplant England (www.nhsbt.nhs.uk), which has supported field work and other elements of the trial. DNA extraction and genotyping were co-funded by the National Institute for Health and Care Research (NIHR), the NIHR BioResource (bioresource.nihr.ac.uk) and the NIHR Cambridge Biomedical Research Centre (BRC-1215-20014) [*]. The academic coordinating centre for INTERVAL was supported by core funding from the: NIHR Blood and Transplant Research Unit in Donor Health and Genomics (NIHR BTRU-2014-10024), NIHR BTRU in Donor Health and Behaviour (NIHR203337), UK Medical Research Council (MR/L003120/1), British Heart Foundation (SP/09/002; RG/13/13/30194; RG/18/13/33946) and NIHR Cambridge BRC (BRC-1215-20014) [*]. A complete list of the investigators and contributors to the INTERVAL trial is provided in reference ^19^. The academic coordinating centre would like to thank the blood donor centre staff and blood donors for participating in the INTERVAL trial. This work was supported by Health Data Research UK, which is funded by the UK Medical Research Council, Engineering and Physical Sciences Research Council, Economic and Social Research Council, Department of Health and Social Care (England), Chief Scientist Office of the Scottish Government Health and Social Care Directorates, Health and Social Care Research and Development Division (Welsh Government), Public Health Agency (Northern Ireland), British Heart Foundation and Wellcome. The authors would like to thank colleagues at the University of Cambridge and the National Haemoglobinopathy Reference Laboratory for their help with preparing and shipping INTERVAL samples and conducting HbF assays. *The views expressed are those of the author(s) and not necessarily those of the NIHR, NHSBT, or the Department of Health and Social Care.

The All of Us Research Program is supported by the National Institutes of Health, Office of the Director: Regional Medical Centers: 1 OT2 OD026549; 1 OT2 OD026554; 1 OT2 OD026557; 1 OT2 OD026556; 1 OT2 OD026550; 1 OT2 OD 026552; 1 OT2 OD026553; 1 OT2 OD026548; 1 OT2 OD026551; 1 OT2 OD026555; IAA #: AOD 16037; Federally Qualified Health Centers: HHSN 263201600085U; Data and Research Center: 5 U2C OD023196; Biobank: 1 U24 OD023121; The Participant Center: U24 OD023176; Participant Technology Systems Center: 1 U24 OD023163; Communications and Engagement: 3 OT2 OD023205; 3 OT2 OD023206; and Community Partners: 1 OT2 OD025277; 3 OT2 OD025315; 1 OT2 OD025337; 1 OT2 OD025276. In addition, the All of Us Research Program would not be possible without the partnership of its participants.

P. Deelen is supported by an NWO ZonMW-VENI Grant (no. 9150161910057).
